# Analysis of Mechanistic Pathways in the Treatment of Non-Alcoholic Steatohepatitis. Evidence from a Bayesian Network Meta-Analysis

**DOI:** 10.1101/2021.10.22.21265361

**Authors:** Mark D. Muthiah, Cheng Han Ng, Jieling Xiao, Yip Han Chin, Grace Lim, Wen Hui Lim, Phoebe Tay, Darren Jun Hao Tan, Jie Ning Yong, Xin-Hui Pan, Jeffery Wei Heng Koh, Nicholas Chew, Nicholas Syn, Eunice Tan, Daniel Q. Huang, Mohammad Shadab Siddiqui, Rohit Loomba, Arun J. Sanyal, Mazen Noureddin

## Abstract

**Background and Aims:** Non-alcoholic steatohepatitis (NASH) is the most common cause of liver disease contributing to significant disease burden worldwide. However, there is lack of comparison of efficacy between different NASH drug classes. We conducted a network meta-analysis evaluating drug classes through comparing histological outcomes and targets of drugs.

**Approach & Results:** Medline, EMBASE and CENTRAL were searched for articles evaluating NASH drugs in biopsy-proven NASH patients. Primary outcomes included NASH resolution without worsening of fibrosis, 2-point reduction in Non-alcoholic fatty liver disease Activity Score (NAS) without worsening of fibrosis and 1-point reduction in fibrosis. Treatments were classified into inflammation, energy, bile acid, and fibrosis modulators. The analysis was conducted with Bayesian network model and surface under the cumulative ranking curve (SUCRA) analysis.

From the 48 trials included, treatments modulating energy (Risk ratio (RR): 1.84, Credible intervals (Crl): 1.29 - 2.65) were the most likely to achieve NASH resolution followed by treatments modulating fibrosis (RR 1.68, Crl: 0.55 - 5.28), bile acids (RR: 1.34, Crl: 0.78 - 2.26) and inflammation (RR: 0.94, Crl: 0.59 - 1.46). Energy and bile acids modulation were effective in 2-point NAS reduction without worsening of fibrosis (RR: 1.60, Crl 1.13 - 2.30 and RR: 1.79, Crl 1.14 - 2.86) and 1-point fibrosis (RR: 1.27, Crl:1.05 - 1.52 and RR: 1.54, Crl: 1.20 - 1.97).

**Conclusions:** This network analysis demonstrates the relative superiority of drugs modulating energy pathways and bile acids in NASH treatment. This guides the development and selection of drugs for combination therapies.

## INTRODUCTION

Non-alcoholic steatohepatitis (NASH) remains the commonest cause of liver disease contributing to a significant burden on the economy in developed countries^1^. NASH is an inflammatory subtype of non-alcoholic fatty liver disease (NAFLD) with the presence of steatosis, hepatocyte injury (ballooning), inflammation, with or without fibrosis. In the United States alone, the prevalence of NASH is estimated to be between 3% - 5% and projected to increase rapidly, mirroring the rise in obesity^2^. Despite its major implications, the Food and Drug Administration (FDA) has yet to approve any pharmacological treatments for NASH.

While multiple drugs have entered phase III trials, limited efficacy at best have been demonstrated^3^. The basis of NASH treatment targets various steps in the aetiological pathway of NASH (Figure 1). Broadly, this can be classified into (1) surplus energy provision to the hepatocytes, (2) hepatocyte death and inflammation, and (3) development of fibrosis and (4) extrahepatic factors that can also modulate the progression of the disease. NASH occurs when there is excess adiposity and energy delivery to the liver. When hepatocytes are unable to cope with the excess energy, they form lipotoxic metabolites^4^. These lipotoxic metabolites give rise to apoptosis, cell death, and inflammation, primarily mediated by mononuclear cells^5^. Chronic ongoing inflammation then leads to fibrosis, with activation of hepatic stellate cells to myofibroblasts and accumulation of the extracellular matrix^6^. While fibrosis is the main feature associated with mortality and liver related events^7^, attempts at reversing this critical aspect of the disease have been dismal^8^. Extrahepatic factors that modify delivery of energy into the liver and systemic insulin resistance can also modulate the progression of the disease.

**Figure 1:**
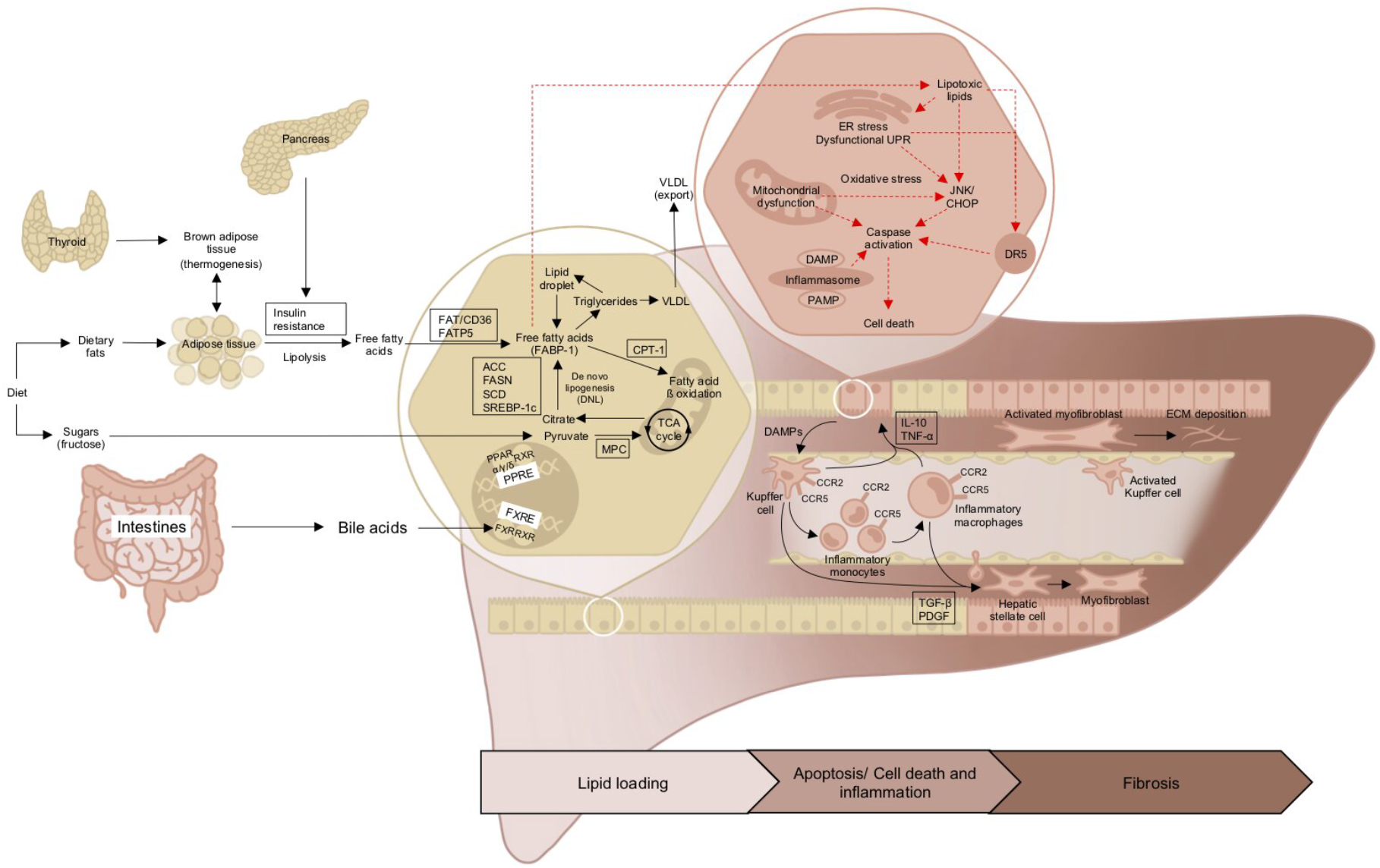
Illustration of basis of non-alcoholic steatohepatitis treatment targets in aetiological pathway of non-alcoholic steatohepatitis

In turn, it is imperative to target key pathways to optimize improvements in outcomes in NASH to serve as the backbone in combination therapies. A previous meta-analysis done by Majzoub et al conducted a surface under the curve cumulative ranking probabilities analysis^11^ and building on this, we sought to examine the comparative efficacy of specific drug classes and targets that can achieve improvements in histological endpoints. To address this knowledge gap, we performed a network meta-analysis comparing the histological outcomes of various classes and targets of drugs used in the treatment of NASH.

## METHODS

### Search Strategy

The present study has been registered with PROSPERO (CRD: CRD42021272676). The network meta-analysis was conducted with reference to the Preferred Reporting Items for Systematic Reviews and Meta-analyses extended statement for network analysis^12, 13^. A search was conducted with assistance from a medical librarian for NASH randomized controlled trials (RCTs) with an updated search conducted on 28^th^ September 2021. Articles was included from inception without the use of a date filter. An example of the search strategy can be found in supplementary material 1. References were managed using Endnote X9 for duplicate removal and references.

### Eligibility and Selection Criteria

Four authors (MDM, CHN, YHC, JX) were involved in the screening of abstracts and evaluation of full text for inclusion based on the eligibility criteria. In this meta-analysis, only English articles which included adult patients with biopsy proven NASH were considered for inclusion. Paediatric studies were excluded. Trials examining a combination of drugs within the same treatment arm were excluded. Based on provided data, the primary outcomes of this meta-analysis were (i) the resolution of NASH without worsening of fibrosis, (ii) 2-point reduction in NAS score without worsening of fibrosis and (iii) at least 1-point reduction in fibrosis. The secondary outcomes of the meta-analysis included an at least a 1-point reduction in steatosis, ballooning and inflammation from liver biopsy. Additionally, biochemical reduction of aspartate aminotransferase (AST) and alanine transaminase (ALT) were outcomes of interest. When articles did not present continuous variables in mean and standard deviations, formulas from Wan et al and author et al were used in the estimation of mean and standard deviations which are required for the pooling of continuous variables. We considered only the use of full text articles, with conference abstracts excluded from the meta anaylsis.

### Classifications of Treatments

The classification of treatments in this meta-analysis is illustrated in Figure 2 and primary articles were grouped into four major groups namely into (1) inflammation (2) energy (3) fibrosis and (4) bile acids based on the mechanism of action.

**Figure 2:**
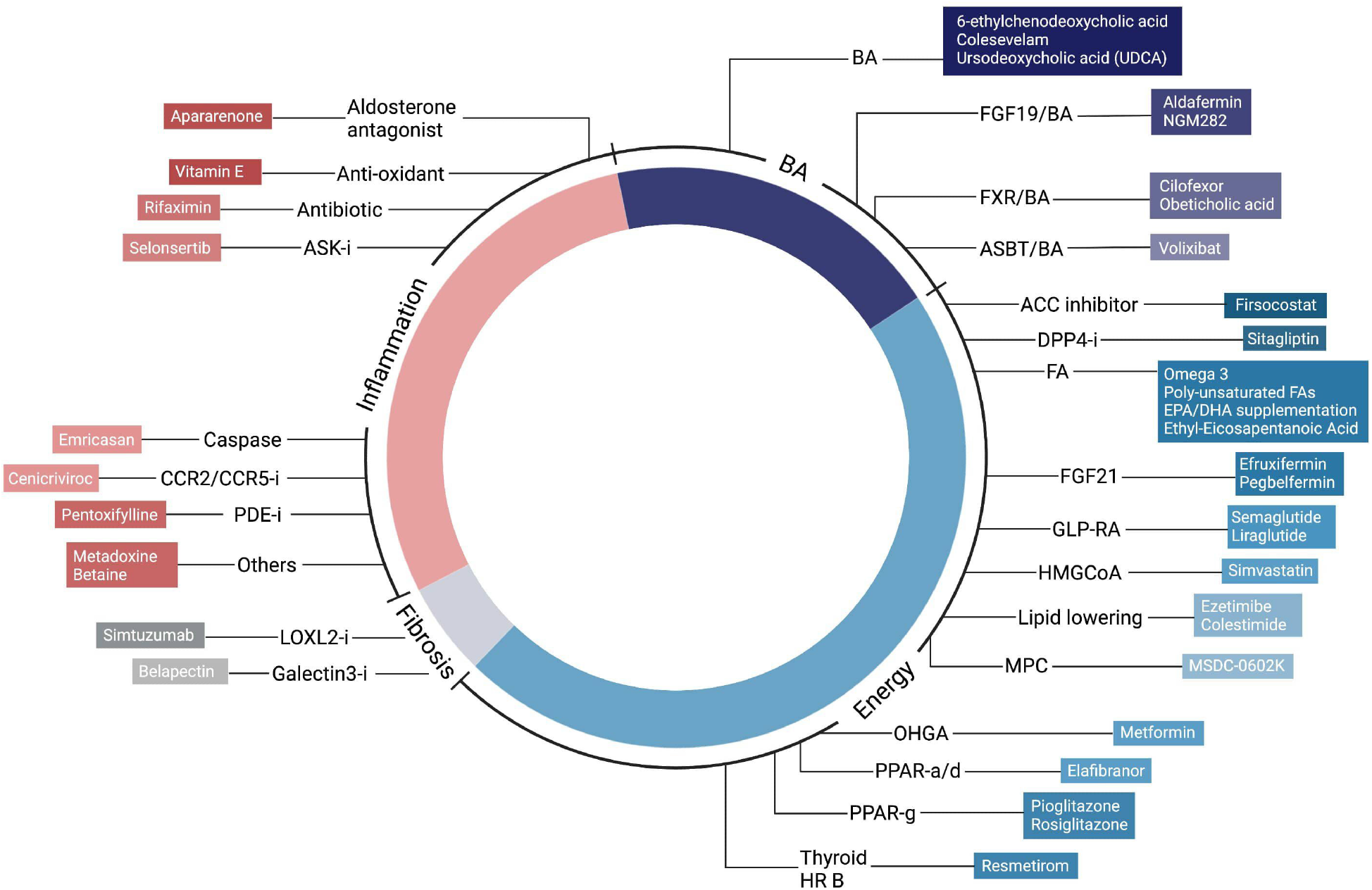
Classification of treatments. (ASK-i: Apoptosis signal-regulating kinase inhibitor, CCR2/CCR5-i: C-C chemokine receptor type 2/ C-C chemokine receptor type 5 inhibitor, PDE-i: Phosphodiesterase inhibitors, LOXL2-i: Lysyl Oxidase Like 2 inhibitor, Galectin3-i: Galectin-3 inhibitors, Thyroid HR B: Thyroid hormone receptor-β, PPAR-g: Peroxisome proliferator-activated receptor gamma, PPAR-a/d: Peroxisome proliferator-activated receptor alpha/delta, OHGA: Oral hypoglycemic agent, MPC: Mitochondrial pyruvate carrier, HMGCoA: 3-hydroxy-3-methylglutaryl-CoA, GLP-RA: Glucagon-like peptide-1 receptor agonists, FGF21: Fibroblast growth factor 21, FAs: Fatty acids, EPA: Eicosapentaenoic acid, DHA: Docosahexaenoic acid, DPP4-i: Dipeptidyl peptidase-4 inhibitors, ACC: Acetyl-CoA carboxylases, ASBT/BA: Apical Sodium Dependent Bile Acid Transporter/bile acid, FXR/BA: farnesoid X receptor/bile acid, FGF19/BA: Fibroblast growth factor 19/bile acid)

### Risk of Bias Assessment

The risk of bias assessment was assessed using the Cochrane Risk of Bias 2.0. Briefly, included articles were examined on seven domains including random sequence generation, allocation concealment, masking of participants and personnel, blinding of outcome assessment, incomplete outcome data, selective outcome reporting, and other sources of bias. Disagreements were resolved by consensus or appeal to a third author.

### Statistical Analysis

Statistical analysis was conducted in RStudio (R version 4.0.3). The analysis was conducted in a Bayesian network model from a generalized liner model using BUGSnet and JAGS software. The unit of measure in the network meta-analysis was risk ratio (RR) for dichotomous events and mean difference (MD) for continuous events with a log-link and identity-link respectively. Bayes iterations parameters were set to 1000 burn-ins, 1000 adaptations, and 10000 iterations for the Markov Chain Monte Carlo (MCMC) algorithm^14^. Model fit was examined from a visual inspection of the trace and density plot. In view of the small sample sizes involve in the primary articles, surface under the curve cumulative ranking probabilities (SUCRA) analysis was considered the endpoint of treatment outcomes. The SUCRA analysis ranks each treatment group from 0-1 with a higher number relating to an increase probability of a successful event. Both models of fix and random effects were conducted, and evaluation of model fitting was based on the Deviance Information Criterion (DIC). Consistency, which assesses statistical agreement between indirect and direct evidence required for validation of the transitivity assumption was examined through DIC and unrelated mean effects (UME) model.^14^ The outputs of the meta-analysis were presented in RR/MD with the corresponding credible intervals (Crl). Publication bias was assessed with visual examination of forest plot for asymmetry.

## RESULTS

### Summary of Included Articles

1,435 articles were retrieved form the initial search strategy, with 1,201 remaining after duplicate removal. After screening of title and abstract, 165 full texts were reviewed, of which 117 articles were excluded. A total of 48 RCTs comprising of 6282 participants were included in this meta-analysis. 3,448 participants were assigned to the experiment group while 2,834 participants were assigned to the control group. There were 16 experiment groups evaluating medications classified under the inflammation subset^15-29^, 25 groups under the energy subset^16, 30-52^, 8 groups under the bile acid subset^52-59^ and 3 groups under the fibrosis subset^60, 61^. Majority of the included RCTs had placebo as the control group with exception of 3 articles with Pioglitazone^19^, Simtzutumab^23^ and vitamin E as control^27^. In the case of trials from the same institutional database analysing the same cohort of participants across multiple publications, the most recent publication was included. Summary of the included articles can be found in supplementary material 2. Majority of RCTs were found to have low to moderate risk of bias in at least half of the domains assessed (Supplementary material 3) and there was no evidence of publication bias (Supplementary material 4).

### Primary Outcomes

#### Resolution of NASH without Worsening of Fibrosis

The summary of results can be found in Table 1 and Figure 3. In total 3639 patients were assessed for resolution of NASH without worsening of fibrosis. Results from the SUCRA analysis ranked treatments modulating energy as the most likely to achieve resolution in NASH without worsening of fibrosis, followed by treatments modulating fibrosis, bile acids, inflammation, and placebo respectively. When compared to placebo, treatments modulating energy resulted in statistical significance in the resolution of NASH without worsening of fibrosis (RR: 1.84, Crl: 1.29 to 2.65). There was no significant difference between treatments modulating fibrosis (RR 1.68, Crl: 0.55 to 5.28), bile acids (RR: 1.34, Crl: 0.78 to 2.26), and inflammation (RR: 0.94, Crl: 0.59 to 1.46) when compared to placebo. Comparing between treatments, energy was statistically superior to those modulating inflammation in the resolution of NASH without worsening of fibrosis (RR: 1.95, Crl: 1.17 to 3.41, table 1).

**Table 1:** Comparison of treatments for Resolution of NASH, 2-Point Reduction in NAS and 1-Point Reduction in Fibrosis.

|  | Energy | Fibrosis | Bile Acid | Inflammation | Placebo |
| --- | --- | --- | --- | --- | --- |
| **Resolution of NASH** |  |  |  |  |  |
| Energy | - | 1.09 (0.33, 3.55) | 1.37 (0.74, 2.65) | 1.95 (1.17, 3.41)* | 1.84 (1.29, 2.65)* |
| Fibrosis | 0.92 (0.28, 3.03) | - | 1.26 (0.37, 4.48) | 1.79 (0.54, 6.20) | 1.68 (0.55, 5.28) |
| Bile Acid | 0.73 (0.38, 1.36) | 0.80 (0.22, 2.71) | - | 1.43 (0.71, 2.86) | 1.34 (0.78, 2.26) |
| Inflammation | 0.51 (0.29, 0.86)* | 0.56 (0.16, 1.84) | 0.70 (0.35, 1.41) | - | 0.94 (0.59, 1.46) |
| Placebo | 0.54 (0.38, 0.77)* | 0.59 (0.19, 1.81) | 0.75 (0.44, 1.29) | 1.07 (0.68, 1.69) | - |
| **2-Point Reduction in NAS without Worsening of Fibrosis** |  |  |  |  |  |
| Energy | - | 1.59 (0.71, 3.58) | 0.89 (0.51, 1.56) | 1.19 (0.71, 1.99) | 1.60 (1.13, 2.30)* |
| Fibrosis | 0.63 (0.28, 1.41) | - | 0.56 (0.24, 1.33) | 0.75 (0.34, 1.64) | 1.01 (0.48, 2.09) |
| Bile Acid | 1.12 (0.64, 1.96) | 1.79 (0.75, 4.24) | - | 1.34 (0.72, 2.46) | 1.79 (1.14, 2.86)* |
| Inflammation | 0.84 (0.50, 1.40) | 1.33 (0.61, 2.97) | 0.75 (0.41, 1.38) | - | 1.34 (0.90, 2.03) |
| Placebo | 0.63 (0.43, 0.89)* | 0.99 (0.48, 2.07) | 0.56 (0.35, 0.88)* | 0.75 (0.49, 1.11) | - |
| **1-Point Reduction in Fibrosis** |  |  |  |  |  |
| Energy | - | 1.39 (0.79, 2.44) | 0.82 (0.61, 1.11) | 1.22 (0.98, 1.52) | 1.27 (1.05, 1.52)* |
| Fibrosis | 0.72 (0.41, 1.26) | - | 0.59 (0.33, 1.06) | 0.88 (0.50, 1.53) | 0.91 (0.53, 1.55) |
| Bile Acid | 1.21 (0.90, 1.65) | 1.69 (0.94, 3.04) | - | 1.48 (1.09, 2.00)* | 1.54 (1.20, 1.97)* |
| Inflammation | 0.82 (0.66, 1.02) | 1.14 (0.65, 2.01) | 0.68 (0.50, 0.91)* | - | 1.04 (0.87, 1.24) |
| Placebo | 0.79 (0.66, 0.95)* | 1.10 (0.65, 1.87) | 0.65 (0.51, 0.83)* | 0.96 (0.81, 1.15) | - |

**Legend:** \* denotes statistical significance, NASH = Non-alcoholic steatohepatitis, NAS = Non-alcoholic fatty liver disease (NAFLD) Activity Score

**Figure 3:**
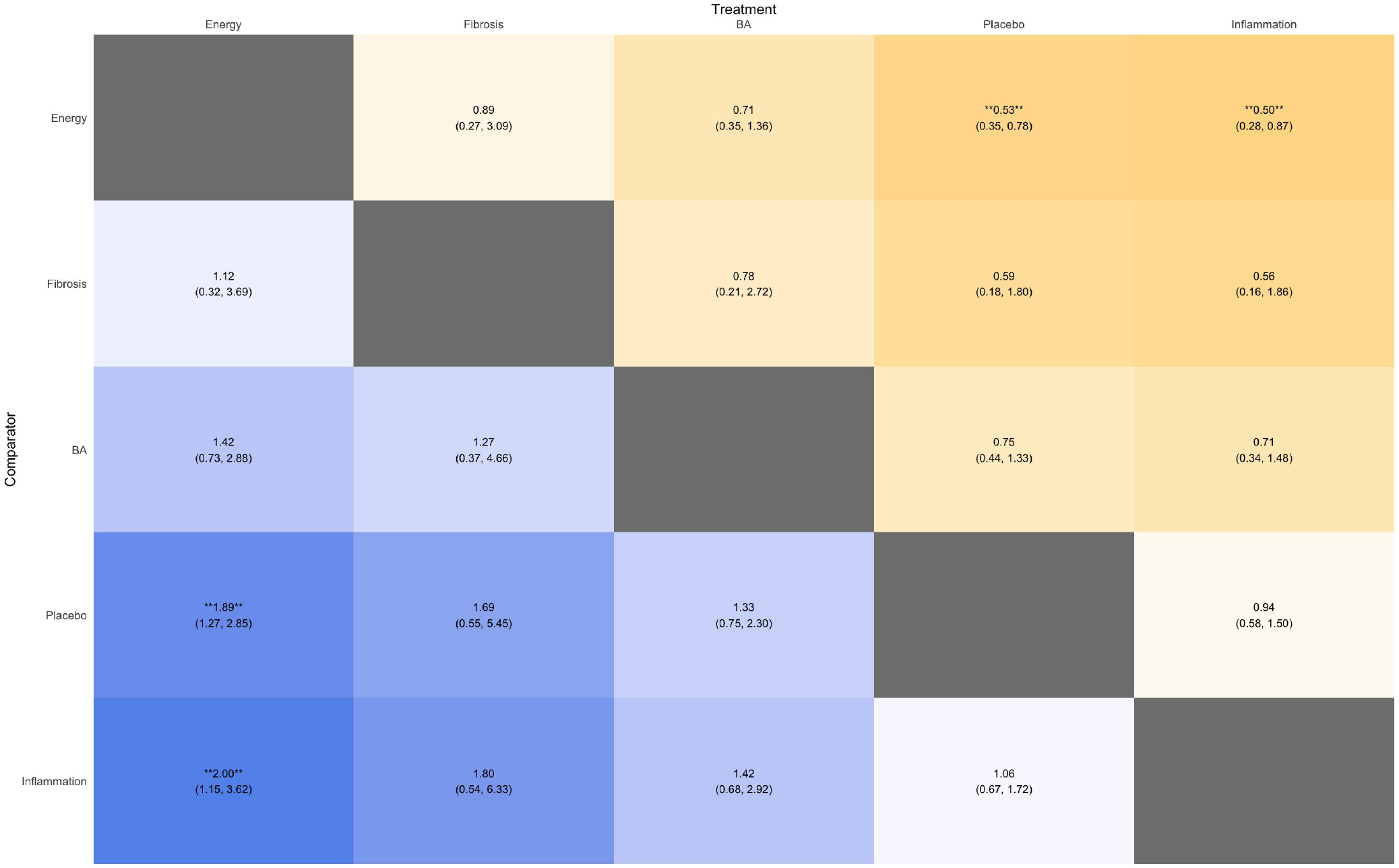
Summary of network heat plot of treatment in resolution of non-alcoholic steatohepatitis without worsening of fibrosis

#### 2-Point Reduction in NAS without Worsening of Fibrosis

There were a total of 3020 patients assessed for a 2-point reduction in NAS without worsening of fibrosis. In the SUCRA analysis, treatments modulating bile acids were ranked as the most likely treatments for a 2-point reduction in NAS without worsening of fibrosis, following which treatments modulating energy, inflammation, fibrosis and placebo respectively were ranked in descending order. Treatments modulating bile acids were statistically more likely than placebo to result in a 2-point reduction of NAS with no worsening of fibrosis (RR: 1.79, Crl 1.14 to 2.86, Table 1). Similarly, treatments modulating energy were significantly better than placebo in a 2-point reduction of NAS without worsening of fibrosis (RR: 1.60, Crl 1.13 to 2.30). There was however no statistical difference when treatments modulating inflammation (RR: 1.34, Crl: 0.90 to 2.03) or fibrosis (RR: 1.01, Crl: 0.48 to 2.09) were compared to placebo. There was no statistically significant difference between groups (Table 1).

#### 1-Point Reduction in Fibrosis

A total of 3568 patients were assessed for a 1-point reduction in fibrosis. SUCRA analysis ranked treatments modulating bile acids as the best treatment in achieving a 1-point reduction in fibrosis, followed by energy, fibrosis, inflammation and placebo respectively. There was statistically significant benefit in treatments modulating bile acids (RR: 1.54, Crl: 1.20 to 1.97) and energy (RR: 1.27, Crl:1.05 to 1.52) when compared to placebo. However, there was no statistical significance between treatments modulating inflammation (RR: 1.04, Crl: 0.87 to 1.24) and fibrosis (RR: 0.91, Crl: 0.53 to 1.55) when compared to placebo. There was no statistically significant difference between groups (Table 1).

### Secondary Outcomes

#### 1-Point Reduction in Steatosis, Ballooning or Lobar Inflammation

The results of the secondary endpoints are summarized in supplementary material 5. A total of 2363 patients were assessed for histological improvements steatosis. In the ranking of treatment, treatments modulating energy were ranked as the best treatment for a histological improvement in steatosis followed by treatments modulating inflammation, bile acids, fibrosis and placebo. There was a significant benefit in 1-point reduction of steatosis between treatments modulating energy when compared to placebo (RR: 1.95, Crl 1.44 to 2.62). Treatments modulating inflammation were also superior to placebo in the reduction of steatosis (RR: 1.64, Crl: 1.20 to 2.26). There was no statistical difference between treatments modulating bile acid (RR: 1.37, Crl: 0.94 to 1.99) or fibrosis (RR: 1.23, Crl: 0.25 to 4.41) when compared to placebo. There was no statistical difference between groups. The results of ballooning and lobar inflammation are summarized in Supplementary table 5. Briefly, both energy and bile acids were statically superior to placebo (RR: 1.40, Crl: 1.04 to 1.91 and RR: 1.56, Crl: 1.04 to 2.34 respectively) with similar results in lobar inflammation.

#### Reduction in Liver Enzymes (AST, ALT)

In total, there was 4874 and 5013 patients examined for AST and ALT respectively. The results of the network analysis are summarized in supplementary material 6. For reduction in AST, treatment ranking in descending order were treatments modulating bile acids, energy, placebo, inflammation, and fibrosis. Treatments modulating bile acids (MD: -7.60, Crl: -11.20 to -3.99) and energy (MD: -6.11, Crl: -6.88 to -5.33) resulted in similar reduction of AST when compared to placebo. There was no statistical difference between treatments modulating inflammation and placebo (MD: 0.68, Crl: -0.81 to 2.18). Treatments modulating fibrosis resulted in an increase in AST compared to placebo (MD: 25.49, Crl: 22.74 – 28.23). Additionally, most treatments were superior to those modulating fibrosis in the reduction of AST. In reduction of ALT, treatments modulating inflammation, bile acids, energy, fibrosis, and placebo were found to be in descending order of treatment ranking. Treatments modulating inflammation resulted in largest reduction in ALT (MD: -6.57, Crl -8.99 to -4.16) when compared to placebo. There was a statistically significant reduction between treatments modulating bile acid (MD: - 5.17, Crl: -7.02 to -3.31) and placebo. Similarly, treatments modulating energy (MD: -3.43, Crl -5.11 to -1.74) and fibrosis (MD: -2.84, -4.19 to -1.50) resulted in a statistically significant reduction in ALT when compared to placebo. Treatments modulating inflammation were superior to most treatments in ALT reduction.

## DISCUSSION

NASH poses a significant burden on the individual, society, and economy^1^. In the absence of efficacious pharmacological treatments for NASH^3^, weight loss remains the only available option for patients to reverse their liver disease^62^. Unfortunately, due to the limited efficacy and sustainability of weight loss, patients are at risk of progressing to end stage complications of the disease that can only be treated by a liver transplant^63^. Current clinical trial design has predominately focus on displaying efficacy compared to placebo treatment. In this class effect network meta-analysis of 48 NASH randomized controlled trials, we demonstrate the comparative benefits of histological endpoints in NASH. Energy modulating treatments were significantly better than placebo in achieving all the 3 primary outcomes (NASH resolution with no worsening of fibrosis, 2-point improvement in NAS with no worsening of fibrosis, and at least 1-point improvement in fibrosis without worsening of steatohepatitis), while bile acid modulating treatments were only significantly better than placebo in achieving 2-point improvement in NAS without worsening if fibrosis and an at least a 1-point improvement in fibrosis without worsening of steatohepatitis (Figure 4). There was no statistical significance between bile acid modulating treatments and placebo in achieving NASH resolution without worsening of fibrosis.

**Figure 4:**
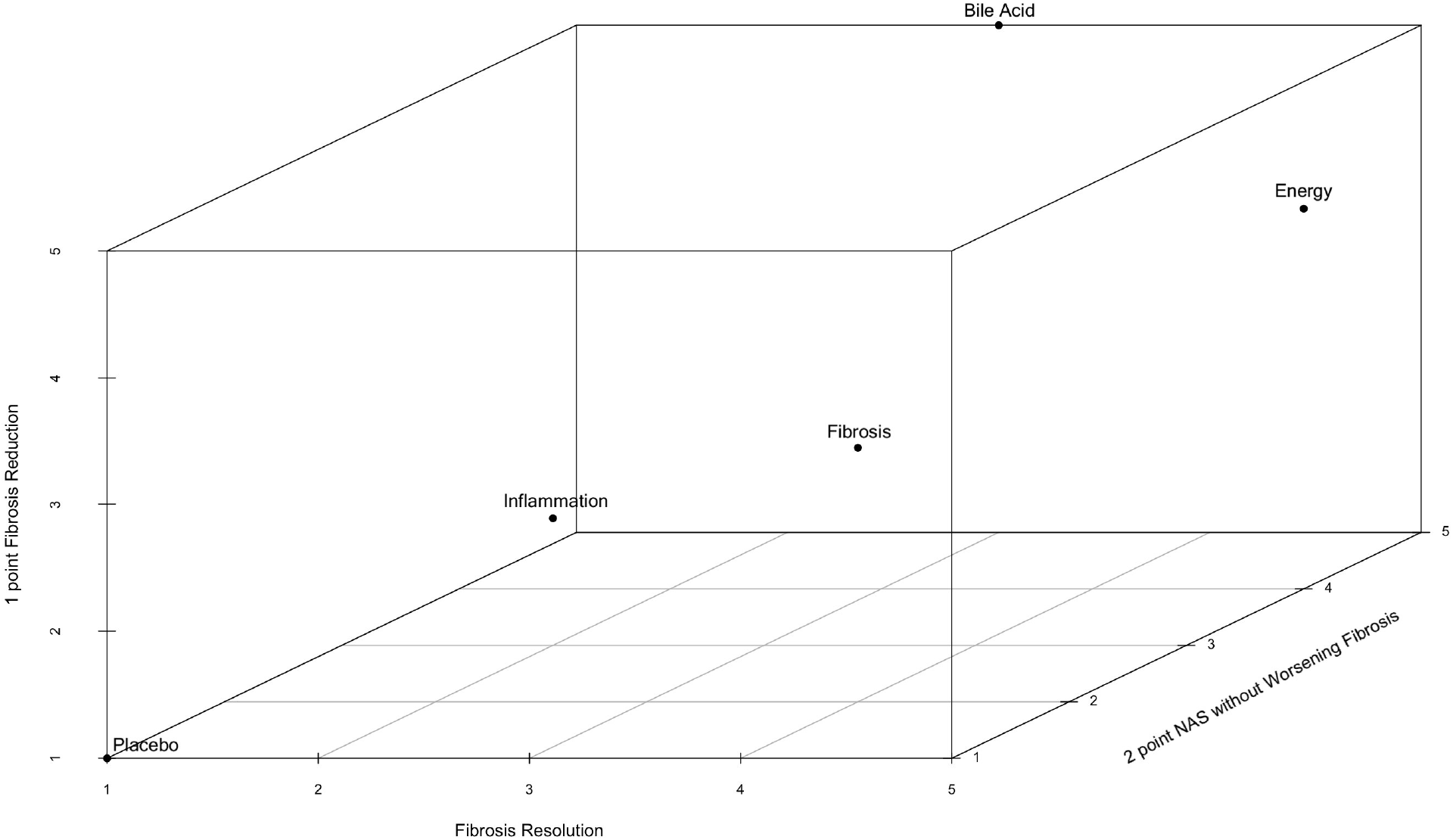
Comparison of drug classes in histological outcomes

In our analysis, treatment modulating energy and bile acids were the most successful treatment for NASH. Within energy modulating treatment, notable studies included the recently published phase 2 trial of subcutaneous semaglutide in which almost half of the patients in the intervention arm achieved resolution of NASH with no worsening of fibrosis. In the PIVENS study, pioglitazone was associated with improvement in hepatic steatosis, lobular inflammation along with serum AST and ALT levels^16^. On the contrary, bile acids are thought to impact both energy delivery into the liver, as well as modulate inflammatory pathways in the liver. Obeticholic acid remains the only NASH drug to achieve a primary endpoint of at least 1-point improvement in fibrosis with no worsening of NASH in a phase 3 study^64^.

While our study demonstrated that both the energy modulating class and the bile acid modulating class appeared to have the best outcomes on liver histology, drugs from these classes reported relevant metabolic adverse effects. Fircostat, an ACC inhibitor from the energy modulating class, can cause hypertriglyceridemia^65^, while obeticholic acid, an FXR agonist from the bile acid modulating class can increase the low-density lipoprotein (LDL) levels^64^. Patients with NASH are at increased risk of cardiovascular disease and cardiovascular related mortality and thus long term results from this adverse effects must also be balanced against the comparative liver related benefits. These drugs may benefit from co-prescriptions with lipid medications. In the phase 2 CONTROL study, obeticholic acid prescribed together with atorvastatin demonstrated safety and tolerability^66^. Evidence from meta-analysis have also shown that statins are safe treatment in NASH^67^. Conversely, some drugs like GLP1-RAs have demonstrated improvements in cardiovascular events and cardiovascular mortality in non-NASH subjects and may have synergistic benefits over and above benefits to just the liver in patients with NASH^68^.

As we only included studies that had paired liver biopsies to demonstrate changes in histology, there were several studies that we were unable to include in the comparisons. PXL770, an AMP-activated kinase (AMPK) activator, and TVB-2640, a fatty acid synthase (FASN) inhibitor, have both demonstrated encouraging results in phase 2a studies^69, 70^. Additionally, combination therapy was also excluded from our analysis. Combination therapy requires principal component analysis to account for a potentiating or addictive effects. Previous literature suggested that combination therapy has the potential to increase response rate from synergistic effects of combined drugs and reduce side effects seen in monotherapy^71^ and remains to be explored.

### Strengths and Limitations

The current analysis provides the first Bayesian network analysis based on the mechanism of action with energy and bile acids modulating treatment showing superiority in achieving the primary endpoint. The basis of the classification of the current network analysis was to achieve a larger sample size for sufficient power to detect significant differences. However, there are several limitations to this meta-analysis. The classification of treatments was based on the proposed pathogenesis of NASH the groupings can potentially be contentious. However, the process was done with expert consensus, and were based around classifications previously described in the field^72^. Additionally, compared to other classes with significantly larger quantity of studies, the recent introduction in anti-fibrotic treatment can be poorly represented. Lastly, current trial design is ill-equipped to assess long term attenuation of outcomes including the rate of cardiovascular events, liver related events, and mortality.

## Conclusion

In conclusion, we demonstrate the relative superiority of drugs modulating energy pathways and bile acids in the treatment of NASH among the available drug targets. These findings further reinforce the hypothesis that the key in treating NASH is not just in trying to improve the pathological changes in the liver, but in targeting the systemic milieu. This serves as a guide to optimize the development of drugs, as well as in selecting drugs for combination therapies. However, more studies are still needed to investigate the longer-term outcomes including both liver related outcomes as well as cardiovascular outcomes in order to justify true benefit to the NASH patient.

## Supporting information

PRISMA 2020 checklist

PRISMA Abstract Checklist

## Data Availability

All articles in this manuscript are publicly available from Medline and Embase

## List of abbreviations

ALT: Alanine transaminase
AMPK: AMP-activated kinase
AST: Aspartate aminotransferase
Crl: Credible intervals
DIC: Deviance Information Criterion
FASN: Fatty Acid Synthase
FDA: Food and Drug Administration
LDL: Low-density Lipoprotein
MCMC: Markov Chain Monte Carlo
MD: Mean Difference
NAFLD: Non-alcoholic Fatty Liver Disease
NAS: Non-alcoholic fatty liver disease (NAFLD) Activity Score
NASH: Non-alcoholic steatohepatitis
RCTs: Randomized Controlled Trials
RR: Risk Ratio
SUCRA: Surface Under the Curve Cumulative Ranking Probabilities
UME: Unrelated Mean Effects

## Acknowledgements

All authors have made substantial contributions to all of the following: (1) the conception and design of the study, or acquisition of data, or analysis and interpretation of data, (2) drafting the article or revising it critically for important intellectual content, (3) final approval of the version to be submitted. No writing assistance was obtained in the preparation of the manuscript. The manuscript, including related data, figures and tables has not been previously published and that the manuscript is not under consideration elsewhere.

## FIGURE AND TABLE LEGENDS

**Table 1:** Comparison of treatments for resolution of non-alcoholic steatohepatitis, 2-Point reduction in NAFLD activity score and 1-Point reduction in fibrosis

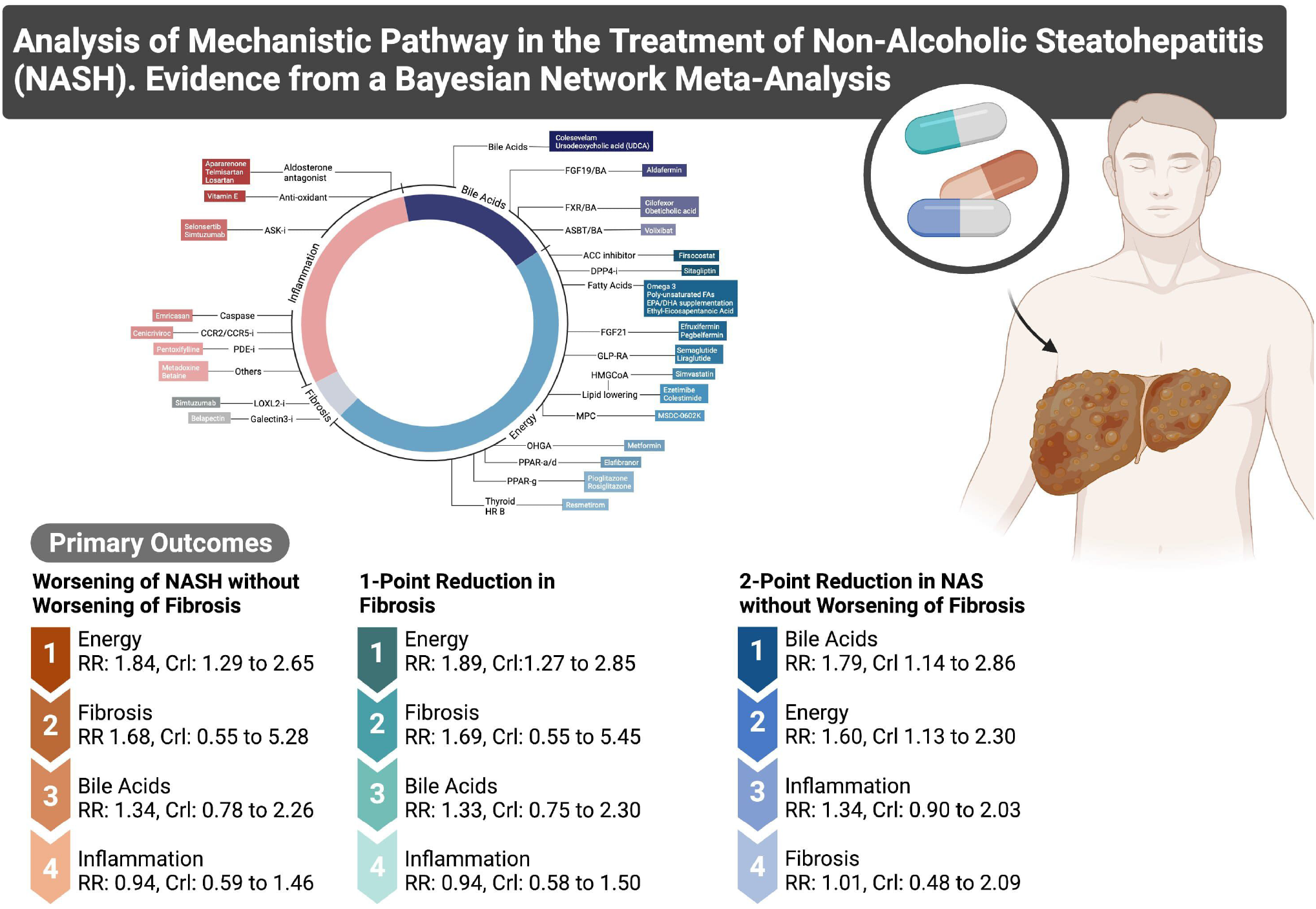

## REFERENCES

1. Muthiah MD, Sanyal AJ. Burden of Disease due to Nonalcoholic Fatty Liver Disease. Gastroenterol Clin North Am. Mar 2020;49(1):1–23. doi:10.1016/j.gtc.2019.09.007

2. Younossi ZM, Koenig AB, Abdelatif D, Fazel Y, Henry L, Wymer M. Global epidemiology of nonalcoholic fatty liver disease-Meta-analytic assessment of prevalence, incidence, and outcomes. Hepatology. 07 2016;64(1):73–84. doi:10.1002/hep.28431

3. Noureddin M, Muthiah MD, Sanyal AJ. Drug discovery and treatment paradigms in nonalcoholic steatohepatitis. Endocrinol Diabetes Metab. Oct 2020;3(4):e00105. doi:10.1002/edm2.105

4. Neuschwander-Tetri BA. Hepatic lipotoxicity and the pathogenesis of nonalcoholic steatohepatitis: the central role of nontriglyceride fatty acid metabolites. Hepatology. Aug 2010;52(2):774–88. doi:10.1002/hep.23719

5. Gadd VL, Skoien R, Powell EE, et al. The portal inflammatory infiltrate and ductular reaction in human nonalcoholic fatty liver disease. Hepatology. Apr 2014;59(4):1393–405. doi:10.1002/hep.26937

6. Puche JE, Saiman Y, Friedman SL. Hepatic stellate cells and liver fibrosis. Compr Physiol. Oct 2013;3(4):1473–92. doi:10.1002/cphy.c120035

7. Angulo P, Kleiner DE, Dam-Larsen S, et al. Liver Fibrosis, but No Other Histologic Features, Is Associated With Long-term Outcomes of Patients With Nonalcoholic Fatty Liver Disease. Gastroenterology. Aug 2015;149(2):389-97.e10. doi:10.1053/j.gastro.2015.04.043

8. Harrison SA, Abdelmalek MF, Caldwell S, et al. Simtuzumab Is Ineffective for Patients With Bridging Fibrosis or Compensated Cirrhosis Caused by Nonalcoholic Steatohepatitis. Gastroenterology. Oct 2018;155(4):1140–1153. doi:10.1053/j.gastro.2018.07.006

9. Marcellin P, Gane E, Buti M, et al. Regression of cirrhosis during treatment with tenofovir disoproxil fumarate for chronic hepatitis B: a 5-year open-label follow-up study. Lancet. Feb 2013;381(9865):468–75. doi:10.1016/S0140-6736(12)61425-1

10. Jang JW, Choi JY, Kim YS, et al. Effects of Virologic Response to Treatment on Short- and Long-term Outcomes of Patients With Chronic Hepatitis B Virus Infection and Decompensated Cirrhosis. Clin Gastroenterol Hepatol. Dec 2018;16(12):1954-1963.e3. doi:10.1016/j.cgh.2018.04.063

11. Majzoub AM, Nayfeh T, Barnard A, et al. Systematic review with network meta-analysis: comparative efficacy of pharmacologic therapies for fibrosis improvement and resolution of NASH. https://doi.org/10.1111/apt.16583. Alimentary Pharmacology & Therapeutics. 2021/10/01 2021;54(7):880-889. doi:https://doi.org/10.1111/apt.16583

12. Page MJ, McKenzie JE, Bossuyt PM, et al. The PRISMA 2020 statement: an updated guideline for reporting systematic reviews. BMJ. 2021;372:n71. doi:10.1136/bmj.n71

13. Hutton B, Salanti G, Caldwell DM, et al. The PRISMA Extension Statement for Reporting of Systematic Reviews Incorporating Network Meta-analyses of Health Care Interventions: Checklist and Explanations. Annals of Internal Medicine. 2015/06/02 2015;162(11):777–784. doi:10.7326/M14-2385

14. Béliveau A, Boyne DJ, Slater J, Brenner D, Arora P. BUGSnet: an R package to facilitate the conduct and reporting of Bayesian network Meta-analyses. BMC Medical Research Methodology. 2019/10/22 2019;19(1):196. doi:10.1186/s12874-019-0829-2

15. Abdelmalek MF, Sanderson SO, Angulo P, et al. Betaine for nonalcoholic fatty liver disease: results of a randomized placebo-controlled trial. Hepatology (Baltimore, Md). 2009;50(6):1818- Erratum in: Hepatology. 2010 May;51(5):1868Comment in: Hepatology. 2010 May;51(5):1859; author reply 1859-60; PMID: 20432267 [https://www.ncbi.nlm.nih.gov/pubmed/20432267]. doi:https://dx.doi.org/10.1002/hep.23239

16. Sanyal AJ, Chalasani N, Kowdley KV, et al. Pioglitazone, vitamin E, or placebo for nonalcoholic steatohepatitis. The New England journal of medicine. 2010;362(18):1675–85. Comment in: Hepatology. 2010 Aug;52(2):789-92; PMID: 20683969 doi:https://dx.doi.org/10.1056/NEJMoa0907929

17. Zein CO, Lopez R, Fu X, et al. Pentoxifylline decreases oxidized lipid products in nonalcoholic steatohepatitis: new evidence on the potential therapeutic mechanism. Hepatology (Baltimore, Md). 2012;56(4):1291–9. doi:https://dx.doi.org/10.1002/hep.25778

18. Van Wagner LB, Koppe SWP, Brunt EM, et al. Pentoxifylline for the treatment of non-alcoholic steatohepatitis: a randomized controlled trial. Annals of hepatology. 2011;10(3):277–86.

19. Sharma BC, Kumar A, Garg V, Reddy RS, Sakhuja P, Sarin SK. A Randomized Controlled Trial Comparing Efficacy of Pentoxifylline and Pioglitazone on Metabolic Factors and Liver Histology in Patients with Non-alcoholic Steatohepatitis. Journal of clinical and experimental hepatology. 2012;2(4):333–7. doi:https://dx.doi.org/10.1016/j.jceh.2012.10.010

20. Shenoy KT, Balakumaran LK, Mathew P, et al. Metadoxine Versus Placebo for the Treatment of Non-alcoholic Steatohepatitis: A Randomized Controlled Trial. Journal of clinical and experimental hepatology. 2014;4(2):94–100. doi:https://dx.doi.org/10.1016/j.jceh.2014.03.041

21. Alam S, Nazmul Hasan S, Mustafa G, Alam M, Kamal M, Ahmad N. Effect of Pentoxifylline on Histological Activity and Fibrosis of Nonalcoholic Steatohepatitis Patients: A One Year Randomized Control Trial. Journal of translational internal medicine. 2017;5(3):155–163. doi:https://dx.doi.org/10.1515/jtim-2017-0021

22. McPherson S, Wilkinson N, Tiniakos D, et al. A randomised controlled trial of losartan as an anti-fibrotic agent in non-alcoholic steatohepatitis. PloS one. 2017;12(4):e0175717. doi:https://dx.doi.org/10.1371/journal.pone.0175717

23. Loomba R, Lawitz E, Mantry PS, et al. The ASK1 inhibitor selonsertib in patients with nonalcoholic steatohepatitis: A randomized, phase 2 trial. Hepatology (Baltimore, Md). 2018;67(2):549–559. Comment in: Nat Rev Gastroenterol Hepatol. 2017 Nov;14(11):631; PMID: 29018270 [https://www.ncbi.nlm.nih.gov/pubmed/29018270]Erratum in: Hepatology. 2018 May;67(5):2063; PMID: 29669401 [https://www.ncbi.nlm.nih.gov/pubmed/29669401]. doi:https://dx.doi.org/10.1002/hep.29514

24. Bril F, Biernacki DM, Kalavalapalli S, et al. Role of Vitamin E for Nonalcoholic Steatohepatitis in Patients With Type 2 Diabetes: A Randomized Controlled Trial. Diabetes care. 2019;42(8):1481–1488. doi:https://dx.doi.org/10.2337/dc19-0167

25. Harrison SA, Goodman Z, Jabbar A, et al. A randomized, placebo-controlled trial of emricasan in patients with NASH and F1-F3 fibrosis. Journal of hepatology. 2020;72(5):816–827. doi:https://dx.doi.org/10.1016/j.jhep.2019.11.024

26. Harrison SA, Wong VW-S, Okanoue T, et al. Selonsertib for patients with bridging fibrosis or compensated cirrhosis due to NASH: Results from randomized phase III STELLAR trials. Journal of hepatology. 2020;73(1):26–39. Comment in: J Hepatol. 2020 Jul;73(1):9-11; PMID: 32360996 [https://www.ncbi.nlm.nih.gov/pubmed/32360996]. doi:https://dx.doi.org/10.1016/j.jhep.2020.02.027

27. Alam S, Abrar M, Islam S, et al. Effect of telmisartan and vitamin E on liver histopathology with non-alcoholic steatohepatitis: A randomized, open-label, noninferiority trial. JGH open : an open access journal of gastroenterology and hepatology. 2020;4(4):663–669. doi:https://dx.doi.org/10.1002/jgh3.12315

28. Ratziu V, Sanyal A, Harrison SA, et al. Cenicriviroc Treatment for Adults With Nonalcoholic Steatohepatitis and Fibrosis: Final Analysis of the Phase 2b CENTAUR Study. Hepatology (Baltimore, Md). 2020;72(3):892–905. doi:https://dx.doi.org/10.1002/hep.31108

29. Okanoue T, Sakamoto M, Harada K, et al. Efficacy and safety of apararenone (MT-3995) in patients with nonalcoholic steatohepatitis: A randomized controlled study. Hepatology research : the official journal of the Japan Society of Hepatology. 2021;doi:https://dx.doi.org/10.1111/hepr.13695

30. Uygun A, Kadayifci A, Isik AT, et al. Metformin in the treatment of patients with non-alcoholic steatohepatitis. Alimentary pharmacology & therapeutics. 2004;19(5):537–44. Comment in: Aliment Pharmacol Ther. 2020 Jan;51(1):199-200; PMID: 31850557 [https://www.ncbi.nlm.nih.gov/pubmed/31850557].

31. Belfort R, Harrison SA, Brown K, et al. A placebo-controlled trial of pioglitazone in subjects with nonalcoholic steatohepatitis. The New England journal of medicine. 2006;355(22):2297–307. Comment in: N Engl J Med. 2006 Nov 30;355(22):2361-3; PMID: 17135591

32. Aithal GP, Thomas JA, Kaye PV, et al. Randomized, placebo-controlled trial of pioglitazone in nondiabetic subjects with nonalcoholic steatohepatitis. Gastroenterology. 2008;135(4):1176–84. Comment in: Ann Intern Med. 2009 Jan 20;150(2):JC1-9; PMID: 19172714 [https://www.ncbi.nlm.nih.gov/pubmed/19172714]. doi:https://dx.doi.org/10.1053/j.gastro.2008.06.047

33. Taniai M, Hashimoto E, Tobari M, et al. Treatment of nonalcoholic steatohepatitis with colestimide. Hepatology research : the official journal of the Japan Society of Hepatology. 2009;39(7):685–93. doi:https://dx.doi.org/10.1111/j.1872-034X.2009.00507.x

34. Ratziu V, Giral P, Jacqueminet S, et al. Rosiglitazone for nonalcoholic steatohepatitis: one-year results of the randomized placebo-controlled Fatty Liver Improvement with Rosiglitazone Therapy (FLIRT) Trial. Gastroenterology. 2008;135(1):100–10. Comment in: Gastroenterology. 2008 Dec;135i6):2156; PMID: 19013165 [https://www.ncbi.nlm.nih.gov/pubmed/19013165]. doi:https://dx.doi.org/10.1053/j.gastro.2008.03.078

35. Shields WW, Thompson KE, Grice GA, Harrison SA, Coyle WJ. The Effect of Metformin and Standard Therapy versus Standard Therapy alone in Nondiabetic Patients with Insulin Resistance and Nonalcoholic Steatohepatitis (NASH): A Pilot Trial. Therapeutic advances in gastroenterology. 2009;2(3):157–63. doi:https://dx.doi.org/10.1177/1756283X09105462

36. Nelson A, Torres DM, Morgan AE, Fincke C, Harrison SA. A pilot study using simvastatin in the treatment of nonalcoholic steatohepatitis: A randomized placebo-controlled trial. Journal of clinical gastroenterology. 2009;43(10):990–4. doi:https://dx.doi.org/10.1097/MCG.0b013e31819c392e

37. Sanyal AJ, Abdelmalek MF, Suzuki A, Cummings OW, Chojkier M, Group E-AS. No significant effects of ethyl-eicosapentanoic acid on histologic features of nonalcoholic steatohepatitis in a phase 2 trial. Gastroenterology. 2014;147(2):377-84.e1. Comment in: Gastroenterology. 2015 Jan;148(1):262-3; PMID: 25451655 [https://www.ncbi.nlm.nih.gov/pubmed/25451655]Comment in: Gastroenterology. 2015 Jan;148(1):262; PMID: 25451662 [https://www.ncbi.nlm.nih.gov/pubmed/25451662]. doi:https://dx.doi.org/10.1053/j.gastro.2014.04.046

38. Loomba R, Sirlin CB, Ang B, et al. Ezetimibe for the treatment of nonalcoholic steatohepatitis: assessment by novel magnetic resonance imaging and magnetic resonance elastography in a randomized trial (MOZART trial). Hepatology (Baltimore, Md). 2015;61(4):1239–50. doi:https://dx.doi.org/10.1002/hep.27647

39. Nogueira MA, Oliveira CP, Ferreira Alves VA, et al. Omega-3 polyunsaturated fatty acids in treating non-alcoholic steatohepatitis: A randomized, double-blind, placebo-controlled trial. Clinical nutrition (Edinburgh, Scotland). 2016;35(3):578–86. doi:https://dx.doi.org/10.1016/j.clnu.2015.05.001

40. Li Y-H, Yang L-H, Sha K-H, Liu T-G, Zhang L-G, Liu X-X. Efficacy of poly-unsaturated fatty acid therapy on patients with nonalcoholic steatohepatitis. World journal of gastroenterology. 2015;21(22):7008–13. doi:https://dx.doi.org/10.3748/wjg.v21.i22.7008

41. Dasarathy S, Dasarathy J, Khiyami A, et al. Double-blind randomized placebo-controlled clinical trial of omega 3 fatty acids for the treatment of diabetic patients with nonalcoholic steatohepatitis. Journal of clinical gastroenterology. 2015;49(2):137–44. Comment in: J Clin Gastroenterol. 2016 Feb;50(2):180; PMID: 26448136 [https://www.ncbi.nlm.nih.gov/pubmed/26448136]Comment in: J Clin Gastroenterol. 2016 Feb;50(2):181; PMID: 26583270 [https://www.ncbi.nlm.nih.gov/pubmed/26583270]. doi:https://dx.doi.org/10.1097/MCG.0000000000000099

42. Argo CK, Patrie JT, Lackner C, et al. Effects of n-3 fish oil on metabolic and histological parameters in NASH: a double-blind, randomized, placebo-controlled trial. Journal of hepatology. 2015;62(1):190–7. doi:https://dx.doi.org/10.1016/j.jhep.2014.08.036

43. Ratziu V, Harrison SA, Francque S, et al. Elafibranor, an Agonist of the Peroxisome Proliferator-Activated Receptor-alpha and -delta, Induces Resolution of Nonalcoholic Steatohepatitis Without Fibrosis Worsening. Gastroenterology. 2016;150(5):1147-1159.e5. doi:https://dx.doi.org/10.1053/j.gastro.2016.01.038

44. Cusi K, Orsak B, Bril F, et al. Long-Term Pioglitazone Treatment for Patients With Nonalcoholic Steatohepatitis and Prediabetes or Type 2 Diabetes Mellitus: A Randomized Trial. Annals of internal medicine. 2016;165(5):305–15. Comment in: Ann Intern Med. 2016 Sep 6;165(5):373-4; PMID: 27322890 doi:https://dx.doi.org/10.7326/M15-1774

45. Armstrong MJ, Gaunt P, Aithal GP, et al. Liraglutide safety and efficacy in patients with non-alcoholic steatohepatitis (LEAN): a multicentre, double-blind, randomised, placebo-controlled phase 2 study. Lancet (London, England). 2016;387(10019):679–90. Comment in: Lancet. 2016 Feb 13;387(10019):628-30; PMID: 26608257 [https://www.ncbi.nlm.nih.gov/pubmed/26608257]Comment in: Lancet. 2016 Jun 11;387(10036):2378-9; PMID: 27312302

46. Joy TR, McKenzie CA, Tirona RG, et al. Sitagliptin in patients with non-alcoholic steatohepatitis: A randomized, placebo-controlled trial. World journal of gastroenterology. 2017;23(1):141–150. doi:https://dx.doi.org/10.3748/wjg.v23.i1.141

47. Alam S, Ghosh J, Mustafa G, Kamal M, Ahmad N. Effect of sitagliptin on hepatic histological activity and fibrosis of nonalcoholic steatohepatitis patients: a 1-year randomized control trial. Hepatic medicine : evidence and research. 2018;10:23–31. doi:https://dx.doi.org/10.2147/HMER.S158053

48. Harrison SA, Ruane PJ, Freilich BL, et al. Efruxifermin in non-alcoholic steatohepatitis: a randomized, double-blind, placebo-controlled, phase 2a trial. Nature medicine. 2021;27(7):1262–1271. doi:https://dx.doi.org/10.1038/s41591-021-01425-3

49. Harrison SA, Alkhouri N, Davison BA, et al. Insulin sensitizer MSDC-0602K in non-alcoholic steatohepatitis: A randomized, double-blind, placebo-controlled phase IIb study. Journal of hepatology. 2020;72(4):613–626. doi:https://dx.doi.org/10.1016/j.jhep.2019.10.023

50. Harrison SA, Bashir MR, Guy CD, et al. Resmetirom (MGL-3196) for the treatment of non-alcoholic steatohepatitis: a multicentre, randomised, double-blind, placebo-controlled, phase 2 trial. Lancet (London, England). 2019;394(10213):2012–2024. Comment in: Lancet. 2019 Nov 30;394(10213):1970-1972; PMID: 31740030 doi:https://dx.doi.org/10.1016/S0140-6736(19)32517-6

51. Huang JF, Dai CY, Huang CF, et al. First-in-Asian double-blind randomized trial to assess the efficacy and safety of insulin sensitizer in nonalcoholic steatohepatitis patients. Hepatol Int. Aug 12 2021;doi:10.1007/s12072-021-10242-2

52. Loomba R, Noureddin M, Kowdley KV, et al. Combination Therapies Including Cilofexor and Firsocostat for Bridging Fibrosis and Cirrhosis Attributable to NASH. Hepatology (Baltimore, Md). 2021;73(2):625–643. doi:https://dx.doi.org/10.1002/hep.31622

53. Lindor KD, Kowdley KV, Heathcote EJ, et al. Ursodeoxycholic acid for treatment of nonalcoholic steatohepatitis: results of a randomized trial. Hepatology (Baltimore, Md). 2004;39(3):770–8. Comment in: Hepatology. 2004 Mar;39(3):602-3; PMID: 14999677 [https://www.ncbi.nlm.nih.gov/pubmed/14999677].

54. Leuschner UFH, Lindenthal B, Herrmann G, et al. High-dose ursodeoxycholic acid therapy for nonalcoholic steatohepatitis: a double-blind, randomized, placebo-controlled trial. Hepatology (Baltimore, Md). 2010;52(2):472–9. doi:https://dx.doi.org/10.1002/hep.23727

55. Le T-A, Chen J, Changchien C, et al. Effect of colesevelam on liver fat quantified by magnetic resonance in nonalcoholic steatohepatitis: a randomized controlled trial. Hepatology (Baltimore, Md). 2012;56(3):922–32. doi:https://dx.doi.org/10.1002/hep.25731

56. Neuschwander-Tetri BA, Loomba R, Sanyal AJ, et al. Farnesoid X nuclear receptor ligand obeticholic acid for non-cirrhotic, non-alcoholic steatohepatitis (FLINT): a multicentre, randomised, placebo-controlled trial. Lancet (London, England). 2015;385(9972):956–65. Erratum in: Lancet. 2015 Mar 14;385(9972):946Comment in: Lancet. 2015 Mar 14;385(9972):922-4; PMID: 25468161 doi:https://dx.doi.org/10.1016/S0140-6736(14)61933-4

57. Newsome PN, Palmer M, Freilich B, et al. Volixibat in adults with non-alcoholic steatohepatitis: 24-week interim analysis from a randomized, phase II study. Journal of hepatology. 2020;73(2):231–240. doi:https://dx.doi.org/10.1016/j.jhep.2020.03.024

58. Younossi ZM, Ratziu V, Loomba R, et al. Obeticholic acid for the treatment of non-alcoholic steatohepatitis: interim analysis from a multicentre, randomised, placebo-controlled phase 3 trial. Lancet (London, England). 2019;394(10215):2184–2196. Comment in: Lancet. 2019 Dec 14;394(10215):2131-2133; PMID: 31813639

59. Harrison SA, Neff G, Guy CD, et al. Efficacy and Safety of Aldafermin, an Engineered FGF19 Analog, in a Randomized, Double-Blind, Placebo-Controlled Trial of Patients With Nonalcoholic Steatohepatitis. Gastroenterology. 2021;160(1):219-231.e1. doi:https://dx.doi.org/10.1053/j.gastro.2020.08.004

60. Harrison SA, Abdelmalek MF, Caldwell S, et al. Simtuzumab Is Ineffective for Patients With Bridging Fibrosis or Compensated Cirrhosis Caused by Nonalcoholic Steatohepatitis. Gastroenterology. 2018;155(4):1140–1153. doi:https://dx.doi.org/10.1053/j.gastro.2018.07.006

61. Chalasani N, Abdelmalek MF, Garcia-Tsao G, et al. Effects of Belapectin, an Inhibitor of Galectin-3, in Patients With Nonalcoholic Steatohepatitis With Cirrhosis and Portal Hypertension. Gastroenterology. Apr 2020;158(5):1334-1345.e5. doi:10.1053/j.gastro.2019.11.296

62. Vilar-Gomez E, Martinez-Perez Y, Calzadilla-Bertot L, et al. Weight Loss Through Lifestyle Modification Significantly Reduces Features of Nonalcoholic Steatohepatitis. Gastroenterology. Aug 2015;149(2):367-78.e5; quiz e14-5. doi:10.1053/j.gastro.2015.04.005

63. Muthiah MD, Sanyal AJ. Current management of non-alcoholic steatohepatitis. Liver Int. Feb 2020;40 Suppl 1(Suppl 1):89–95. doi:10.1111/liv.14355

64. Younossi ZM, Ratziu V, Loomba R, et al. Obeticholic acid for the treatment of non-alcoholic steatohepatitis: interim analysis from a multicentre, randomised, placebo-controlled phase 3 trial. Lancet. Dec 14 2019;394(10215):2184–2196. doi:10.1016/s0140-6736(19)33041-7

65. Kim CW, Addy C, Kusunoki J, et al. Acetyl CoA Carboxylase Inhibition Reduces Hepatic Steatosis but Elevates Plasma Triglycerides in Mice and Humans: A Bedside to Bench Investigation. Cell Metab. Aug 1 2017;26(2):394-406.e6. doi:10.1016/j.cmet.2017.07.009

66. Pockros PJ, Fuchs M, Freilich B, et al. CONTROL: A randomized phase 2 study of obeticholic acid and atorvastatin on lipoproteins in nonalcoholic steatohepatitis patients. Liver Int. Nov 2019;39(11):2082–2093. doi:10.1111/liv.14209

67. Eslami L, Merat S, Malekzadeh R, Nasseri-Moghaddam S, Aramin H. Statins for non-alcoholic fatty liver disease and non-alcoholic steatohepatitis. Cochrane Database Syst Rev. Dec 27 2013;(12):Cd008623. doi:10.1002/14651858.CD008623.pub2

68. Bethel MA, Patel RA, Merrill P, et al. Cardiovascular outcomes with glucagon-like peptide-1 receptor agonists in patients with type 2 diabetes: a meta-analysis. Lancet Diabetes Endocrinol. Feb 2018;6(2):105–113. doi:10.1016/s2213-8587(17)30412-6

69. Loomba R, Mohseni R, Lucas KJ, et al. TVB-2640 (FASN Inhibitor) for the Treatment of Nonalcoholic Steatohepatitis: FASCINATE-1, a Randomized, Placebo-Controlled Phase 2a Trial. Gastroenterology. Jul 23 2021;doi:10.1053/j.gastro.2021.07.025

70. Cusi K, Alkhouri N, Harrison SA, et al. Efficacy and safety of PXL770, a direct AMP kinase activator, for the treatment of non-alcoholic fatty liver disease (STAMP-NAFLD): a randomised, double-blind, placebo-controlled, phase 2a study. Lancet Gastroenterol Hepatol. Nov 2021;6(11):889–902. doi:10.1016/s2468-1253(21)00300-9

71. Dufour JF, Caussy C, Loomba R. Combination therapy for non-alcoholic steatohepatitis: rationale, opportunities and challenges. Gut. Oct 2020;69(10):1877–1884. doi:10.1136/gutjnl-2019-319104

72. Ratziu V. A critical review of endpoints for non-cirrhotic NASH therapeutic trials. J Hepatol. Feb 2018;68(2):353–361. doi:10.1016/j.jhep.2017.12.001

