## Supplementary Material 1 for "Analysis of Mechanistic Pathways in the Treatment of Non-Alcoholic Steatohepatitis. Evidence from a Bayesian Network Meta-Analysis"

**Supplementary material 1: Search Strategy**

1. (clinical adj3 trial).tw. or (singl$ OR doubl$ OR trebl$ OR tripl$).tw. AND (mask$ OR blind$).tw. or placebo$.tw. or random$.tw. or exp randomized controlled trials/ or exp random allocation/ or exp double-blind method/ or exp single-blind method/ or exp placebos/ or research design/

2. (nash or ((nonalcoholic or non*alcoholic or non alcoholic) adj3 steatohepatit*)).tw.
