## Supplementary Material 2 for "Analysis of Mechanistic Pathways in the Treatment of Non-Alcoholic Steatohepatitis. Evidence from a Bayesian Network Meta-Analysis"

**Supplementary material 2: Summary of Included Articles**

| Author (Year) | Country | Medication (Experiment) | | Medication (Control) | Age (years) | | Male (no./total sample size) | | BMI | | Risk of Bias |
| --- | --- | --- | --- | --- | --- | --- | --- | --- | --- | --- | --- |
|  |  | Medication | Subset | Medication | Experiment | Control | Experiment | Control | Experiment | Control |  |
| Uygun et al (2004) | Turkey | Metformin 1700 mg/day | Energy | Placebo | 39.8 (10.6) | 41.5 (9.1) | 11/17 | 10/17 | 30.1 (3.4) | 28.4 (3.9) | High Concern |
| Lindor et al (2004) | Canada | Ursodeoxycholic Acid 13 15mg/kg/day | BA | Placebo | 45.4 (12) | 48.5 (11.6) | 36/80 | 37/86 | 32.3 (6.4) | 31.7 (5.5) | Low Concern |
| Belfort et al (2006) | USA | Pioglitazone 45 mg/day | Energy | Placebo | 51 (7) | 51 (10) | 14/26 | 7/21 | 33.5 (4.9) | 32.9 (4.4) | Low Concern |
| Aithal et al (2008) | UK | Pioglitazone 30 mg/day | Energy | Placebo | 52 (28 - 71)^ | 55 (27 - 73)^ | 26/37 | 19/37 | 29.8 (3.0) | 30.8 (4.1) | Low Concern |
| Taniai et al (2009) | Japan | Colestimide 3g/day | Energy | Placebo | 46 (22.45) | 47 (26.89) | 11/17 | 13/21 | 25.9 (3.97) | 26.68 (4.28) | Some Concerns |
| Ratziu et al (2008) | France | Rosiglitazone (4mg/day for 1 month; 8mg thereafter) | Energy | Placebo | 53.1 (11.5) | 54.1 (10.4) | 19/32 | 18/31 | 31.5 (6) | 30.5 (4.4) | Low Concern |
| Shields et al (2009) | USA | Metfomin 1000mg/day | Energy | Placebo | 50.2 (9.1) | 44.4 (12) | 8/9 | 5/10 | 32.2 (4.9) | 32.8 (4.9) | Some Concerns |
| Nelson et al (2009) | USA | Simvastatin 40 mg/day | Energy | Placebo | 52.6 (8.6) | 52..5 (13) | 7/10 | 4/6 | 37.3 | 34.0 | High Concern |
| Abdelmalek et al (2009) | USA | Betaine 20g/day | Inflammation | Placebo | 47.8 (11.4) | 45.7 (13.4) | 12/27 | 8/28 | 32.4 (6.0) | 34.7 (5.6) | Some Concerns |
| Sanyal et al (2010) | USA | Pioglitazone 30 mg/day | Energy | Placebo | 47.0 (12.6) | 45.4 (11.2) | 21/80 | 25/83 | 34 (6) | 35 (7) | Low Concern |
|  |  | Vitamin E 800IU/day | Inflammation |  | 46.6 (12.1) |  | 22/84 |  | 34 (7) |  | Low Concern |
| Leuschner et al (2010) | Germany and Greece | UDCA 23 - 28 mg/kg/day | BA | Placebo | 41.45 (18 - 71)^ | 45.02 (18 - 73)^ | 63/95 | 63/91 | - | - |  |
| Zein et al (2011) | USA | Pentoxifylline 1200 mg/day | Inflammation | Placebo | 50.5 (12.7) | 49.6 (9.6) | 18/26 | 20/29 | 32.9 (4.6) | 34.0 (5.4) | Some Concerns |
| Van Wagner et al (2011) | USA | Pentoxifylline 1200 mg/day | Inflammation | Placebo | 48 (2) | 53 (2) | 8/21 | 6/9 | 34.0 (0.9) | 35.1(2.6) | Some Concerns |
| Sharma et al (2012) | India | Pentoxifylline 1200 mg/day | Inflammation | Pioglitazone (30 mg/day) | 37.3 (7.2) | 40.4 (9.9) | 7/11 | 4/9 | 24.2 (2.7) | 25.7 (3.58) | Some Concerns |
| Le et al (2012) | USA | Colesevelam 3.75 g/day | BA | Placebo | 45.4 (12.7) | 50.3 (10.4) | 10/25 | 13/25 | 31.3 (4.7) | 31.2 (5.1) | Low Concern |
| Shenoy et al (2014) | India | Metadoxine 1000 mg/day | Inflammation | Placebo | 39.8 (10.4) | 41.1 (8.5) | 52/75 | 43/59 | 26.8 (3.8) | 27.0 (3.9) | Some Concerns |
| Sanyal et al (2014) | North America | Ethyl-Eicosapentanoic Acid 2700 mg/day | Energy | Placebo | 47.8 (11.1) | 50.5 (12.5) | 29/86 | 32/75 | 35.0 (6.3) | 33.6 (5.9) | Some Concerns |
| Loomba et al (2014) | USA | Ezetimibe 10mg/day | Energy | Placebo | 49.0 (14.9) | 49.5 (13.7) | 11/25 | 8/25 | 33.8 (5.2) | 32.9 (5.1) | Low Concern |
| Nogueira et al (2015) | Brazil | Omega 3 acid 1.8mg/day | Energy | Placebo | 52.5 (7.2) | 53.9 (6.8) | 5/32 | 6/28 | 31.1 (4.6) | 30.3 (4.4) | Some Concerns |
| Neuschwander et al (2015) | USA | Obeticholic acid 25mg/day | BA | Placebo | 52 (11) | 51 (12) | 43/141 | 53/142 | 35 (7) | 34 (6) | Low Concern |
| Li et al (2015) | China | PUFA (EPA DHA 1:1 ratio) | Energy | Placebo | 52.6 (6.6) | 50.4 (7.2) | 34/39 | 36/39 | 28.0 (1.4) | 27.2 (1.3) | Some Concerns |
| Dasarathy et al (2015) | USA | PUFA (EPA 2160 mg and DHA 1440 mg) | Energy | Placebo | 51.5 (6.9) | 49.8 (12.1) | 6/18 | 2/19 | 34.8 (4.6) | 35.7 (7.0) | Some Concerns |
| Argo et al (2015) | USA | Omega 3 acid 3000 mg/day | Energy | Placebo | 46.4 (12.1) | 47.2 (12) | 6/17 | 7/17 | 33.3 (8.0) | 31.6 (6.7) | Some Concerns |
| Ratziu et al (2016) | US, Europe | Elafibranor 120 mg/day | Energy | Placebo | 52.4 (11.6) | 52.4 (11.9) | 47/89 | 55/92 | 31.0 (4.4) | 30.9 (4.2) | Some Concerns |
| Cusi et al (2016) | USA | Pioglitazone 30 mg/day (titrated after 2 months to 45 mg/day) | Energy | Placebo | 52 (10) | 49 (11) | 36/50 | 35/51 | 34.3 (4.8) | 34.5 (4.8) | Some Concerns |
| Armstrong et al (2016) | UK | Liraglutide 1.8mg/day | Energy | Placebo | 50 (11) | 52 (12) | 18/26 | 13/26 | 34.2 (4.7) | 37·7 (6·2) | Low Concern |
| Alam et al (2017) | Bangeladesh | Pentoxifylline 1200 mg/day | Inflammation | Placebo | 41.52 (9.85) | 38.80 (6.18) | 7/25 | 5/10 | 27.97 (3.33) | 24.33 (1.48) | Some Concerns |
| Joy et al (2017) | Canada | Sitagliptin 100mg/day | Energy | Placebo | 56.7 (9.9) | 54.7 (9.8) | 3/6 | 2/6 | 35.9 (6.6) | 37.4 (4.7) | Some Concerns |
| McPherson et al (2017) | UK | Losartan 50mg/day | Inflammation | Placebo | 58 (25 - 75)* | 45 (21 - 76)* | 13/24 | 12/21 | 32.8 (26.1 - 43.4)* | 34.1 (26.5 - 45.2)* | Some Concerns |
| Alam et al (2018) | India | Sitagliptin 100 mg/day | Energy | Placebo | 41.7 (9.1) | 35.5 (6.9) | 4/20 | 8/20 | 27.6 (5.1) | 25.3 (2.8) | Some Concerns |
| Abdel-Razik et al (2018) | Egypt | Rifaximin 1100 mg/day | Inflammation | Placebo | 40.2 (9.88) | 38.4 (9.21) | 9/25 | 7/25 | 33.3 (7.45) | 32.8 (7.35) | Some Concerns |
| Harrison et al (2018) | USA | Simtuzumab 125mg/day | Fibrosis | Placebo | 55 (48 - 59)* | 56 (48 - 59)* | 27/74 | 26/74 | 34.1 (30.6 - 38.1)* | 32.7 (29.0 - 37.3)* | Some Concerns |
|  |  | Simtuzumab 700mg/day | Fibrosis | Placebo | 55 (49 - 61)* | 57 (51 - 61)* | 32/86 | 29/85 | 33.1 (29.7 - 37.8)* | 33.8 (29.8 - 38.0)* |  |
| Harrison et al (2018) | USA | NGM282 3mg/day | BA | Placebo | 56.4 (7.8) | 52.8 (11.3) | 12/28 | 7/27 | 34.7 (5.5) | 35.6 (5.8) | Low Concern |
| Loomba et al (2018) | USA | Selonsertib 18mg/day with or without Simtuzumab | Inflammation | Simtzutumab 125mg | 55 (49 - 61)* | 57 (56 - 59)* | 10/32 | 4/10 | 33 (30 - 37)* | 37 (31 - 37)* | Low Concern |
| Portillo-Sanchez et al (2018) | USA | Pioglitazone 30mg/day to 45mg/day | Energy | Placebo | 52 (1) | 50 (2) | 33/46 | 32/46 | 34.2 (0.7) | 34.7 (0.7) | Low Concern |
| Sanyal et al (2018) | USA | Pegbelfermin (BMS-986036) 10mg/day | Energy | Placebo | 52 (10) | 47 (12) | 10/25 | 10/26 | 34 (4) | 37 (7) | Some Concerns |
| Bril et al (2019) | USA | Vitamin E 45mg/day | Inflammation | Placebo | 60 (9) | 57 (11) | 33/36 | 30/32 | 33.8 (4.6) | 33.6 (4.0) | Low Concern |
| Harrison et al (2019) | USA | Efruxifermin 70mg/day | Energy | Placebo | 53.0 (13.2) | 52.4 (9.6) | 9/20 | 6/21 | 37.2 (5.5) | 37.6 (4.8) | Some Concerns |
| Harrison et al (2019) | USA | MSDC-0602K 250mg/day | Energy | Placebo | 56.8 (10.42) | 54.6 (11.21) | 43/101 | 43/94 | 35.03 (6.118) | 35.03 (5.574) | Low Concern |
| Harrison et al (2019) | USA | Resmetirom (MGL-3196) 80mg/day | Energy | Placebo | 51.8 (10·4) | 47.3 (11.7) | 38/84 | 24/41 | 35.8 (6.2) | 33.6 (5.8) | Low Concern |
| Harrison et al (2019) | USA | Emricasan 10mg/day | Inflammation | Placebo | 53.6 (12.0) | 54.6 (11.41) | 89/213 | 53/105 | Median: 34.05 | Median: 33.9 | Low Concern |
| Harrison et al (STELLAR 3) (2019) | USA | Selonsertib 18 mg/day | Inflammation | Placebo | 59 (51 - 64)* | 59 (51 - 63)* | 141/322 | 83/159 | 32.4 (29.5 - 37.1)* | 32.2 (27.5 - 37.5)* | Low Concern |
|  | USA | Selonsertib 18 mg/day | Inflammation | Placebo | 59 (53 - 66)* | 61 (55 - 67)* | 138/354 | 71/172 | 32.4 (28.5 - 37.4)* | 32.9 (27.9 - 37.5)* |  |
| Alam et al (2020) | Bangeladesh | Telmisartan 40 mg/day | Inflammation | Vitamin E 800iu | 36.6 (7.8) | 39.5 (9) | 5/16 | 5/14 | 28.9 (5.4) | 28.8 (3.5) | Some Concerns |
| Chalasani et al (2020) | USA | Belapectin 8 mg/kg | Fibrosis | Placebo | 57.1 (9.3) | 58.4 (8.5) | 11/54 | 18/54 | 34.4 (5.7) | 34.6 (7.1) | Low Concern |
| Newsome et al (2020) | Multinational | Volixibat 5mg/day | BA | Placebo | 52.8 (14.13) | 53.4 (11.75) | 24/49 | 17/49 | - | - | Some Concerns |
| Ratziu et al (2020) | Multinational | Cenicriviroc 150mg/day | Inflammation | Placebo | 54.6 (10.2) | 53.7 (11.0) | 72/145 | 65/144 | 33.6 (5.7) | 34.1 (7.2) | Some Concerns |
| Siddiqui et al (2020) | USA | Obeticholic acid 25mg/day | BA | Placebo | 52 (11) | 50 (12) | 30/99 | 35/97 | 35 (6) | 34 (6) | Some Concerns |
| Younossi et al (2021) | Multinational | Obeticholic acid 25mg/day | BA | Placebo | 55 (11) | 55 (12) | 133/308 | 124/311 | - | - | Low Concern |
| Huang et al (2021) | Taiwan | Pioglitazone 30mg/day | Energy | Placebo | 43.9 (13.7) | 43.8 (11.9) | 27/43 | 39/47 | 28.4 (2.8) | 29.4 (4.6) | Some Concerns |
| Patel et al (2021) | USA | Cilofexor 100mg/day | BA | Placebo | 58 (48 - 64)* | 51 (45 - 56)* | 21/56 | 13/28 | 33.4 (30.3 - 36.6)* | 34.2 (31.5 - 36.0)* | Low Concern |
| Harrison et al (2021) | USA | Aldafermin 1mg/day | BA | Placebo | 53.0 (12.1) | 54.1 (9.7) | 27/53 | 9/25 | 35.8 (6.3) | 36.8 (9.0) | Some Concerns |
| Loomba et al (2021) | USA | Firsocostat 20mg/day | Energy | Placebo | 63 (57 - 68)* | 59 (55 - 66)* | 15/40 | 12/39 | 33 (29 - 39)* | 35 (32 - 39)* | Some Concerns |
|  | USA | Cilofexor 30 mg/day | BA |  | 59 (52 - 65)* |  | 11/40 |  | 32 (29 - 41)* |  | Low Concern |
| Newsome et al (2021) | Multinational | Semaglutide 0.4mg/day | Energy | Placebo | 54.3 (10.2) | 52.4 (10.8) | 35/82 | 36/80 | 35.2 (6.6) | 36.1 (6.6) | Low Concern |
| Okanoue et al (2021) | Japan | Apararenone (MT‐3995) 10mg/day | Inflammation | Placebo | 52 (40 - 67)^ | 53 (31 - 75)^ | 12/24 | 13/23 | 30.2 (21.7 - 39.8)^ | 29.3 (21.8 - 36.3)^ | Some Concerns |

Legend: Values are given in mean and standard deviation unless otherwise stated. * - Values given in median and interquartile range, ^ - Values given in median and range

BA – Bile Acid; BMI – Body mass index; PUFA – Polyunsaturated fatty acid; EPA – Eicosapentaenoic acid; DHA – Docosahexaenoic acid
