## Supplementary Material 3 for "Analysis of Mechanistic Pathways in the Treatment of Non-Alcoholic Steatohepatitis. Evidence from a Bayesian Network Meta-Analysis"

| **Supplementary material 3: Risk of Bias** | | | | | | | |
| --- | --- | --- | --- | --- | --- | --- | --- |
|  | D1 | D2 | D3 | D4 | D5 | D6 | **Overall** |
| Ratziu 2016 | 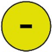 | 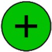 | 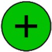 | 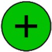 | 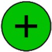 | 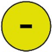 | 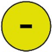 |
| Nogueira 2015 | 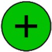 | 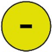 | 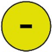 | 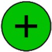 | 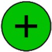 | 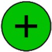 | 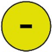 |
| Cusi 2016 | 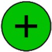 | 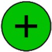 | 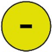 | 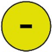 | 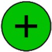 | 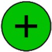 | 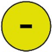 |
| Armstrong 2016 | 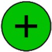 | 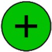 | 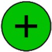 | 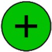 | 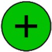 | 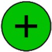 | 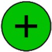 |
| Neuschwander 2015 | 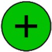 | 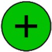 | 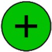 | 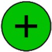 | 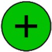 | 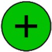 | 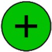 |
| Loomba 2014 | 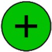 | 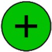 | 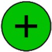 | 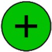 | 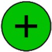 | 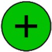 | 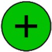 |
| Li 2015 | 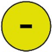 | 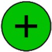 | 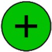 | 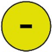 | 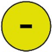 | 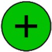 | 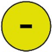 |
| Dasarathy 2015 | 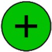 | 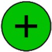 | 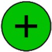 | 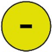 | 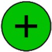 | 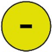 | 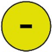 |
| Argo 2015 | 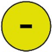 | 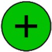 | 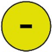 | 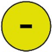 | 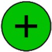 | 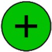 | 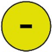 |
| Shenoy 2014 | 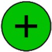 | 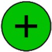 | 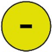 | 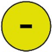 | 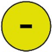 | 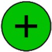 | 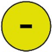 |
| Sanyal 2014 | 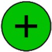 | 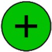 | 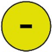 | 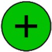 | 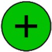 | 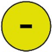 | 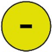 |
| Sharma 2012 | 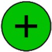 | 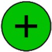 | 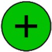 | 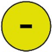 | 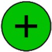 | 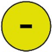 | 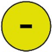 |
| Le 2012 | 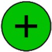 | 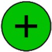 | 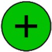 | 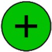 | 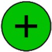 | 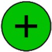 | 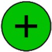 |
| Zein 2011 | 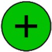 | 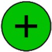 | 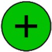 | 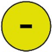 | 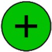 | 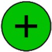 | 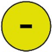 |
| Van Wagner 2011 | 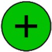 | 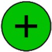 |  |  |  |  |  |
| Sanyal 2010 |  |  |  |  |  |  |  |
| Leuschner 2010 |  |  |  |  |  |  |  |
| Taniai 2009 |  |  |  |  |  |  |  |
| Shields 2009 |  |  |  |  |  |  |  |
| Nelson 2009 |  |  |  |  |  |  |  |
| Abdelmalek 2009 |  |  |  |  |  |  |  |
| Ratziu 2008 |  |  |  |  |  |  |  |
| Aithal 2008 |  |  |  |  |  |  |  |
| Belfort 2006 |  |  |  |  |  |  |  |
| Uygun 2004 |  |  |  |  |  |  |  |
| Lindor 2004 |  |  |  |  |  |  |  |
| Alam 2020 |  |  |  |  |  |  |  |
| Alam 2018 |  |  |  |  |  |  |  |
| Alam 2017 |  |  |  |  |  |  |  |
| Bril 2019 |  |  |  |  |  |  |  |
| Harrison 2018 (Simtuzumab) |  |  |  |  |  |  |  |
| Harrison 2019 (MSDC-0602K) |  |  |  |  |  |  |  |
| Harrison 2019 (MGL-3196 ) |  |  |  |  |  |  |  |
| Harrison 2019 (Emricasan) |  |  |  |  |  |  |  |
| Harrison 2021(Aldafermin) |  |  |  |  |  |  |  |
| Harrison 2019 (Efruxifermin) |  |  |  |  |  |  |  |
| Harrison 2019 (STELLAR) |  |  |  |  |  |  |  |
| Joy 2017 |  |  |  |  |  |  |  |
| Loomba 2018 |  |  |  |  |  |  |  |
| Loomba 2021 |  |  |  |  |  |  |  |
| McPherson 2017 |  |  |  |  |  |  |  |
| Newsome 2021 |  |  |  |  |  |  |  |
| Newsome 2020 |  |  |  |  |  |  |  |
| Okanoue 2021 |  |  |  |  |  |  |  |
| Ratziu 2020 |  |  |  |  |  |  |  |
| Younossi 2021 |  |  |  |  |  |  |  |
| Huang 2021 |  |  |  |  |  |  |  |
| Chalasani 2020 |  |  |  |  |  |  |  |

**Judgement:**

Low concern

Some concerns

High concerns

**Domains:**

D1: Bias arising from the randomization process

D2: Bias due to deviations from intended interventions

D3: Bias due to missing outcome data

D4: Bias in measurement of the outcome

D5: Bias in selection of reported results

D6: Other sources of bias
