## Supplementary Material 4 for "Analysis of Mechanistic Pathways in the Treatment of Non-Alcoholic Steatohepatitis. Evidence from a Bayesian Network Meta-Analysis"

**Supplementary material 4: Funnel Plot for Publication Bias**

**Resolution of NASH without Worsening of Fibrosis**

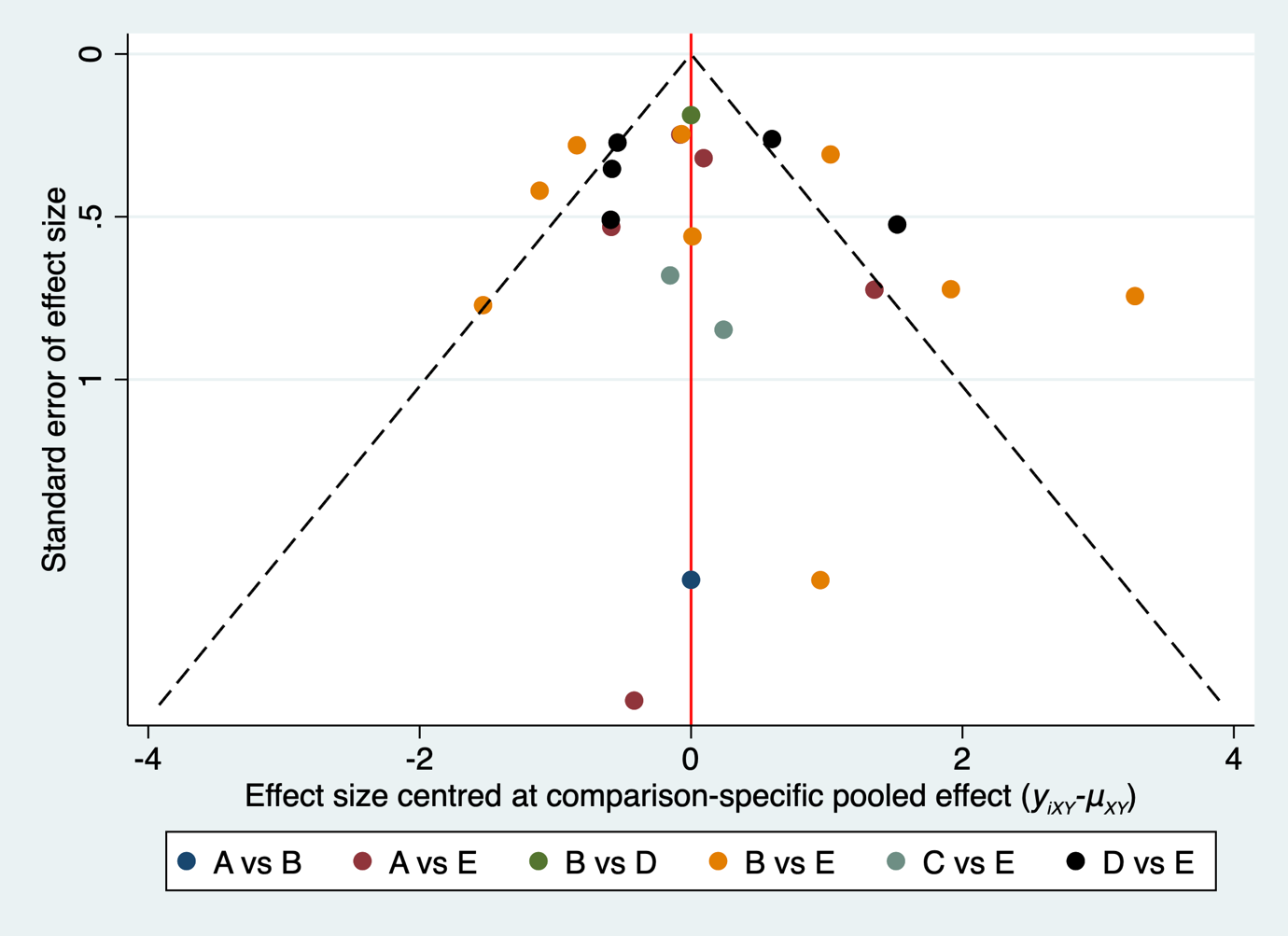

**2-Point Reduction in NAS without Worsening of Fibrosis**

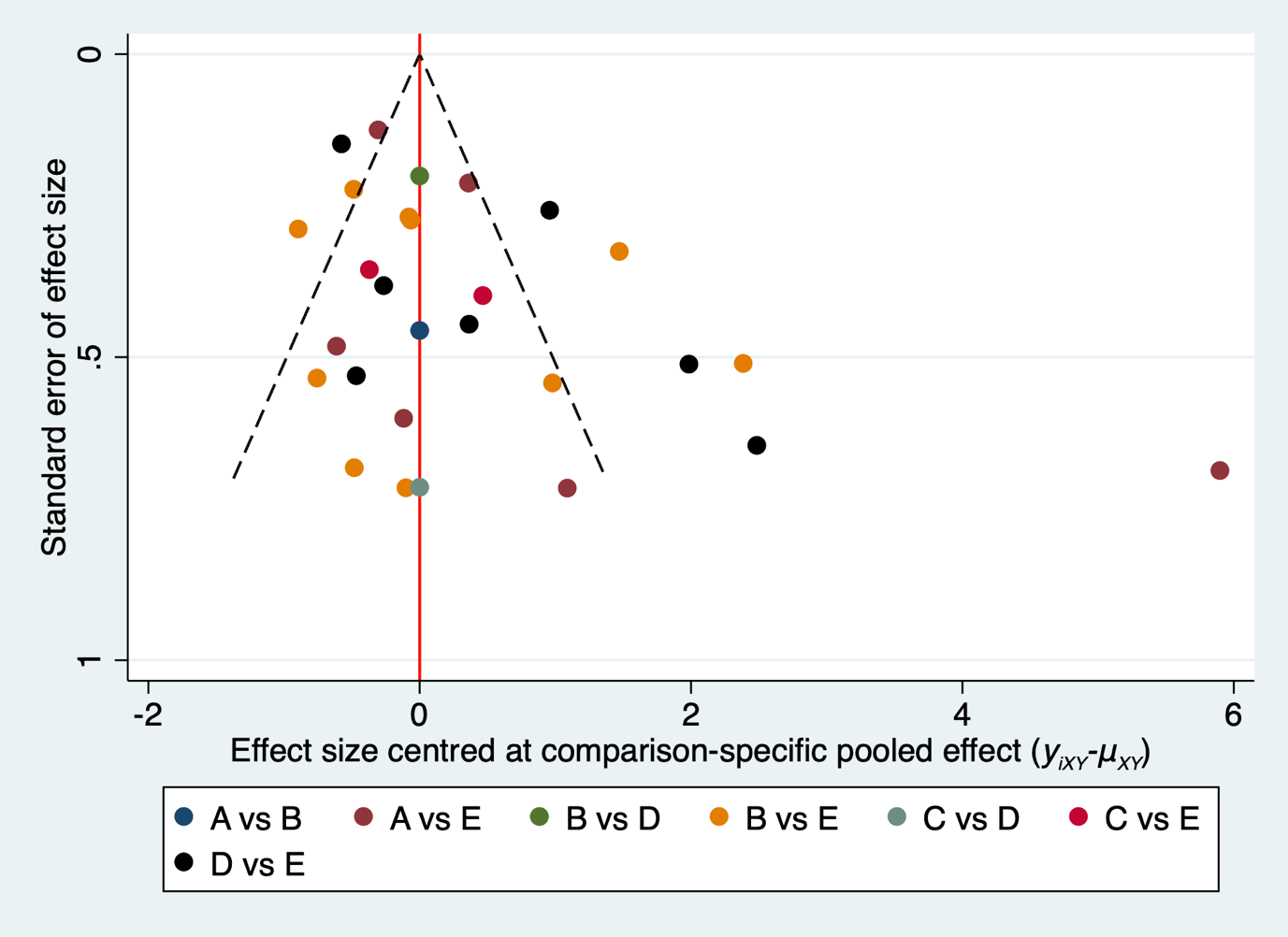

**1-Point Reduction in Fibrosis**

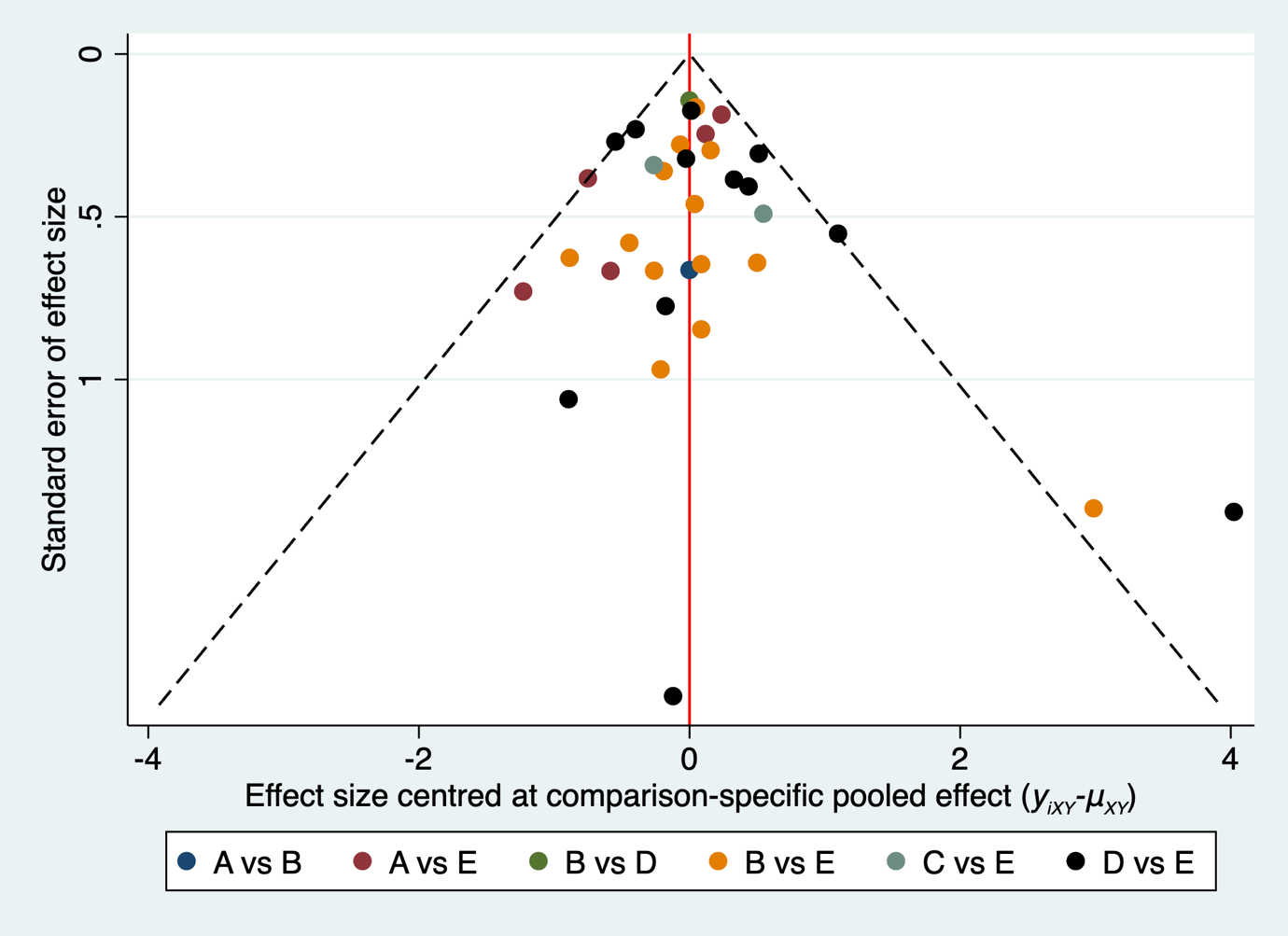
