## Supplementary Material 5 for "Analysis of Mechanistic Pathways in the Treatment of Non-Alcoholic Steatohepatitis. Evidence from a Bayesian Network Meta-Analysis"

**Supplementary material 5: Comparison of Treatments for 1-Point Reduction in Steatosis, Ballooning and Lobar inflammation**

|  | **Energy** | **Fibrosis** | **Bile Acid** | **Inflammation** | **Placebo** |
| --- | --- | --- | --- | --- | --- |
| **1-Point Reduction in Steatosis** | |  |  |  |  |
| Energy |  | 1.58 (0.43, 7.83) | 1.43 (0.89, 2.26) | 1.19 (0.79, 1.77) | 1.95 (1.44, 2.62)* |
| Fibrosis | 0.63 (0.13, 2.32) |  | 0.90 (0.18, 3.38) | 0.75 (0.16, 2.58) | 1.23 (0.25, 4.41) |
| Bile Acid | 0.70 (0.44, 1.12) | 1.11 (0.30, 5.64) | - | 0.84 (0.51, 1.35) | 1.37 (0.94, 1.99) |
| Inflammation | 0.84 (0.57, 1.27) | 1.33 (0.39, 6.31) | 1.20 (0.74, 1.96) | - | 1.64 (1.20, 2.26)* |
| Placebo | 0.51 (0.38, 0.69)* | 0.81 (0.23, 3.96) | 0.73 (0.50, 1.06) | 0.61 (0.44, 0.83)* | - |
| **1-Point Reduction in Ballooning** | | |  |  |  |
| Energy |  | 0.62 (0.14, 2.79) | 0.90 (1.55, 1.47) | 1.32 (0.87, 2.05) | 1.40 (1.04, 1.91)* |
| Fibrosis | 1.62 (0.36, 6.98) |  | 1.46 (0.31, 6.50) | 2.16 (0.50, 8.76) | 2.27 (0.51, 9.56) |
| Bile Acid | 1.12 (0.68, 1.82) | 0.69 (0.15, 3.23) | - | 1.48 (0.87, 2.53) | 1.56 (1.04, 2.34)* |
| Inflammation | 0.76 (0.49, 1.16) | 0.46 (0.11, 2.00) | 0.68 (0.40, 1.15) | - | 1.05 (0.74, 1.49) |
| Placebo | 0.72 (0.52, 0.97)* | 0.44 (0.10, 1.97) | 0.64 (0.43, 0.96)* | 0.95 (0.67, 1.34) | - |
| **1-Point Reduction in Lobar inflammation** | | |  |  |  |
| Energy |  | 1.67 (0.60, 7.06) | 1.14 (0.93, 1.39) | 1.28 (1.09, 1.53)* | 1.47 (1.28, 1.69)* |
| Fibrosis | 0.60 (0.14, 1.66) | - ­ | 0.68 (0.16, 1.92) | 0.77 (0.18, 2.11) | 0.88 (0.21, 2.45) |
| Bile Acid | 0.88 (0.72, 1.07) | 1.47 (0.52, 6.21) | - | 1.13 (0.90, 1.41) | 1.29 (1.11, 1.50)* |
| Inflammation | 0.78 (0.66, 0.92)* | 1.30 (0.47, 5.44) | 0.89 (0.71, 1.11) | - | 1.14 (0.97, 1.35) |
| Placebo | 0.68 (0.59, 0.78)* | 1.14 (0.41, 4.81) | 0.78 (0.67, 0.90)* | 0.87 (0.74, 1.03) | - |

**Legend**: * denotes statistical significance
