## Supplementary Material 6 for "Analysis of Mechanistic Pathways in the Treatment of Non-Alcoholic Steatohepatitis. Evidence from a Bayesian Network Meta-Analysis"

**Supplementary material 6: Comparison of Treatments for Reduction in ALT and AST**

|  | **Energy** | **Fibrosis** | **Bile Acid** | **Inflammation** | **Placebo** |
| --- | --- | --- | --- | --- | --- |
| **Reduction in ALT** | |  |  |  |  |
| Energy | - | -0.58 (-2.73, 1.56) | 1.74 (-0.42, 3.90) | 3.15 (0.21, 6.10)* | -3.43 (-5.11, -1.74)* |
| Fibrosis | 0.58 (-1.56, 2.73) |  | 2.32 (0.02, 4.61)* | 3.73 (0.96, 6.49)* | -2.84 (-4.19, -1.50)* |
| Bile Acid | -1.74 (-3.90, 0.42) | -2.32 (-4.61, -0.02)* | - | 1.41 (-1.64, 4.45) | -5.17 (-7.02, -3.31)* |
| Inflammation | -3.15 (-6.10, -0.21)* | -3.73 (-6.49, -0.98)* | -1.41 (-4.45, 1.64) | - | -6.57 (-8.99, -4.16)* |
| Placebo | 3.43 (1.74, 5.11)* | 2.84 (1.50, 4.19)* | 5.17 (3.31, 7.02)* | 6.57 (4.16, 8.99)* | - |
| **Reduction in AST** | |  |  |  |  |
| Energy |  | -31.60 (-34.45, -28.74)* | 1.49 (-2.20, 5.18) | -6.79 (-8.47, -5.11)* | -6.11 (-6.88, -5.33)* |
| Fibrosis | 31.60 (28.74, 34.45)* |  | 33.09 (28.54, 37.62)* | 24.81 (21.70, 27.91)* | 25.49 (22.74, 28.23)* |
| Bile Acid | -1.49 (-5.18, 2.20) | -33.09 (-37.62, -28.54)* | - | -8.27 (-12.19, -4.37)* | -7.60 (-11.20, -3.99)* |
| Inflammation | 6.79 (5.11, 8.47)* | -24.81 (-27.91, -21.70)* | 8.27 (4.37, 12.19)* | - | 0.68 (-0.81, 2.18) |
| Placebo | 6.11 (5.33, 6.88)* | -25.49 (-28.23, -22.74)* | 7.60 (3.99, 11.20)* | -0.68 (-2.18, 0.81) | - |

**Legend**: * denotes statistical significance, ALT = alanine transaminase, AST = aspartate aminotransferase
